# Monitoring Report: Respiratory Viruses - September 2024 Data

**DOI:** 10.1101/2024.01.20.24301528

**Authors:** Samuel Gratzl, Brianna M Goodwin Cartwright, Patricia J Rodriguez, Charlotte Baker, Duy Do, Nicholas L Stucky

## Abstract

**Background:** Few sources regularly monitor hospitalizations associated with respiratory viruses. This study provides current hospitalization trends associated with six common respiratory viruses: COVID-19, influenza, human metapneumovirus (HMPV), parainfluenza virus, respiratory syncytial virus (RSV), and rhinovirus.

**Objective:** This study aims to supplement the surveillance data provided by the CDC by describing latest trends (through October 13, 2024) overall and for each respiratory virus. This study also provides valuable insight into two at-risk populations: infants and children (age 0-4) and older adults (age 65 and over).

**Methods:** Using a subset of real-world electronic health record (EHR) data from Truveta, a growing collective of health systems that provides more than 18% of all daily clinical care in the US, we identified people who were hospitalized between October 01, 2019 and October 13, 2024. We identified people who tested positive for any of the six respiratory viruses within 14 days of the hospitalization. We report weekly trends in the rate of hospitalizations associated with each virus per all hospitalizations for the overall population and the two high-risk sub populations: infants and children and older adults.

**Results:** We included 748,659 hospitalizations of 692,435 unique patients who tested positive for a respiratory virus between October 01, 2019 and October 13, 2024.

Overall, the rate of hospitalizations associated with respiratory viruses decreased throughout September 2024; there was a 27.9% reduction in the rate of hospitalizations from the first week of September to the first week of October. COVID-associated hospitalizations saw a notable decline of 46.1%, while influenza-associated hospitalizations remained stable. In contrast, there were increases in hospitalizations for parainfluenza virus (+132.6%), RSV (+149.9%), and rhinovirus (+22.8%). In the first week of October 2024, respiratory virus-associated hospitalizations accounted for 2.0% of all hospitalizations. Overall, test positivity rates also decreased, with an overall drop of 39.3%, particularly in COVID positivity (-53.7%). However, rhinovirus test positivity increased slightly to 21.0%, reflecting a 4.5% rise.

**Discussion:** The data indicates a continued decline in respiratory virus-associated hospitalizations across the overall population, particularly for COVID-19. The notable increases in parainfluenza and RSV hospitalizations suggest a shift in the respiratory virus landscape, highlighting the need for ongoing surveillance. The decrease in test positivity rates for COVID-19 is encouraging, but the rising trend in rhinovirus positivity warrants attention as it may indicate a growing prevalence of this virus.

**Trends in surveillance:** *Overall population:* 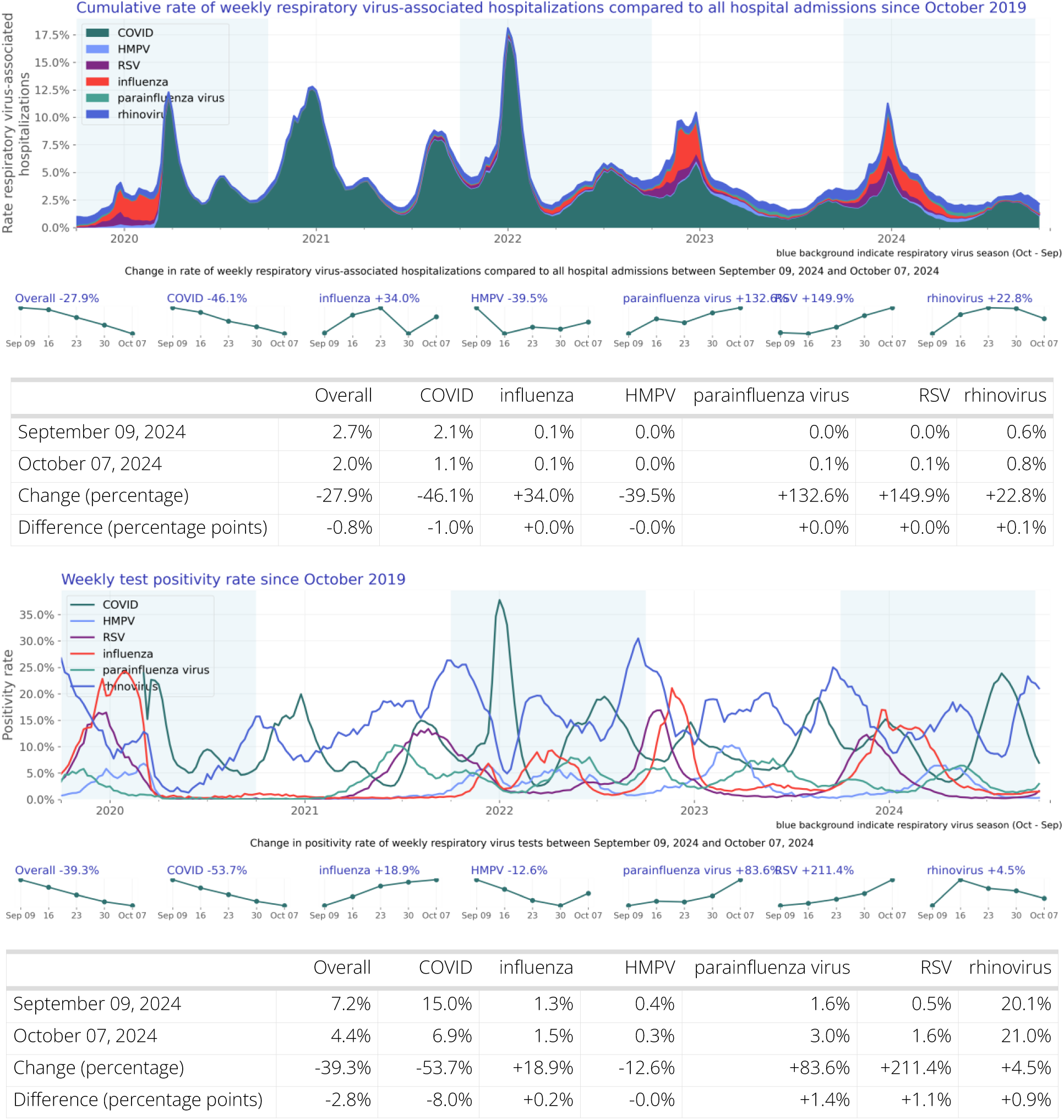 For the overall population, the rate of hospitalizations associated with respiratory viruses decreased by 27.9% in the first week of October 2024 compared to September 2024. The decline in COVID-associated hospitalizations (-46.1%) was substantial, while influenza-associated hospitalizations remained unchanged. Notably, parainfluenza and RSV-associated hospitalizations saw substantial increases of 132.6% and 149.9%, respectively, alongside a smaller increase in rhinovirus-associated hospitalizations (+22.8%). In the first week of October, respiratory virus-associated hospitalizations accounted for 2.0% of all hospitalizations. The overall rate of positive tests decreased by 39.3%, with COVID test positivity dropping substantially (-53.7%). However, rhinovirus test positivity increased slightly to 21.0%, indicating a potential rise in this virus’s prevalence.

*Infants and children (age 0-4):* 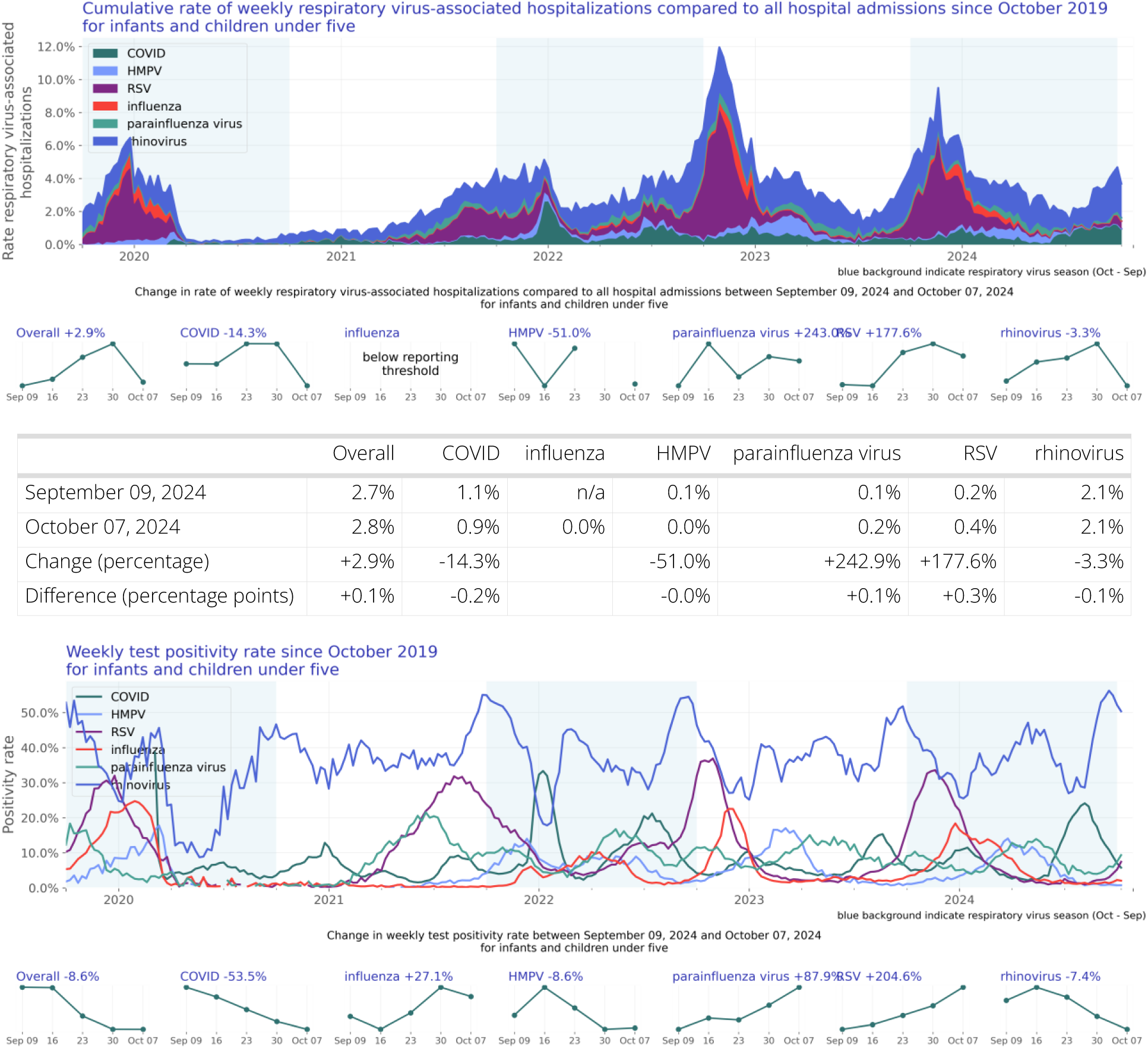

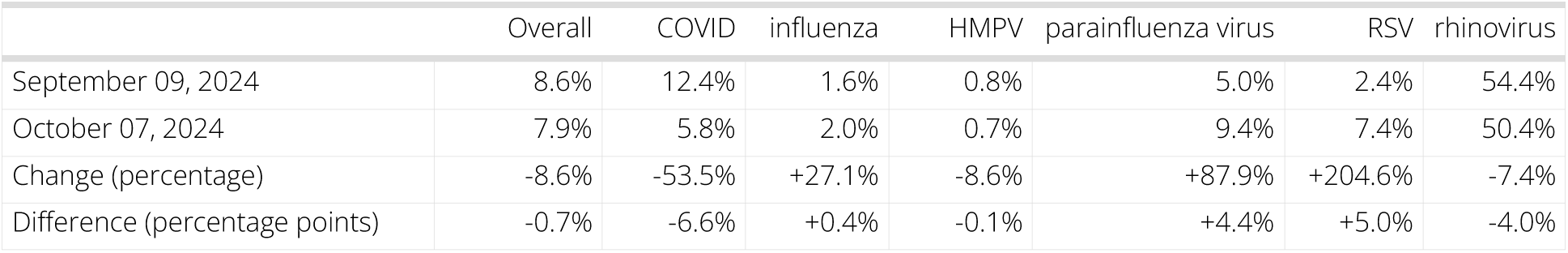 In the population aged 0-4 years, respiratory virus-associated hospitalizations increased slightly by 2.9% throughout September 2024. COVID-associated hospitalizations decreased by 14.3%, while parainfluenza-associated hospitalizations rose drastically (+242.9%) and RSV-associated hospitalizations increased by 177.6%. In the first week of October, 2.8% of all hospitalizations in this age group were associated with a respiratory virus, with rhinovirus being the largest contributor at 2.1%. The overall test positivity rate decreased by 8.6%, with COVID test positivity dropping substantially (-53.5%). Rhinovirus test positivity remained high at 50.4%, although it decreased by 7.4% compared to September.

*Older adults (age 65 and over):* 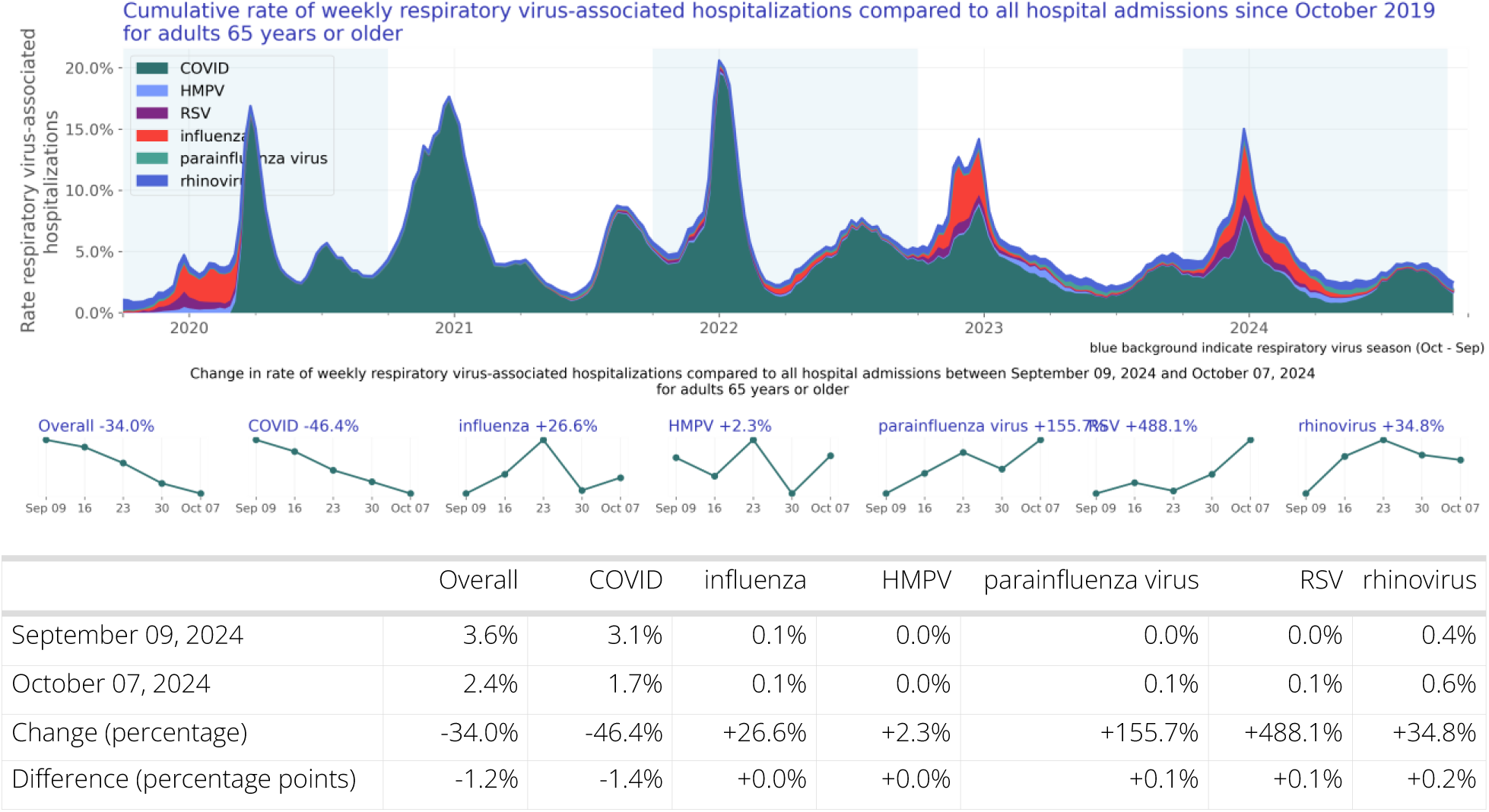

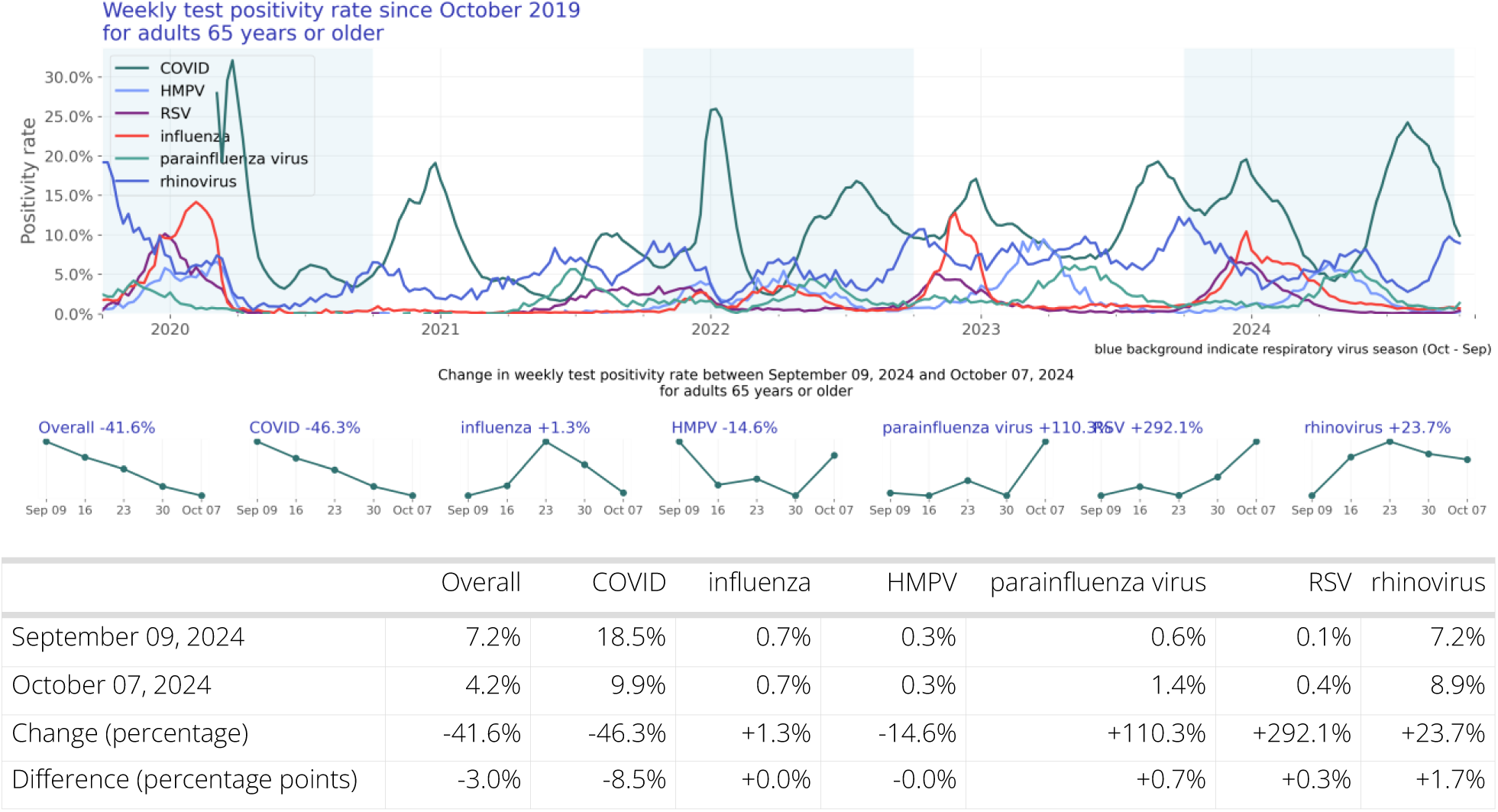 Among older adults (age 65 and over), there was a decrease of 34.0% in hospitalizations associated with respiratory viruses throughout September 2024. COVID-associated hospitalizations decreased by 46.4%, while influenza-associated hospitalizations remained stable. In the first week of October, respiratory virus-associated hospitalizations accounted for 2.4% of all hospitalizations in this age group, with rhinovirus being a notable contributor at 0.6%. The overall rate of positive tests decreased by 41.6%, with COVID test positivity dropping substantially (-46.3%). Rhinovirus test positivity increased to 8.9%, reflecting a 23.7% rise, which may indicate a growing concern for this virus among older adults.

## Introduction

COVID, influenza, and RSV account for a large proportion of hospitalizations related to respiratory illnesses in the United States. To provide a more complete understanding of hospitalizations related to respiratory viruses, we have also included other viruses known to cause respiratory illness such as human metapneumovirus (HMPV), parainfluenza, and rhinovirus. Each of these viruses can lead to hospitalization and death especially in vulnerable populations, such as infants, children, and older adults (Pastula et al., 2017; Shi et al., 2017, Centers for Disease Control and Prevention 2023a, Smits et al., 2023). Representative and timely data to proactively monitor infections are scarce.

Respiratory viruses are also a major source of infection and hospitalizations in older adults (defined here as patients 65 years of age or older). Incidence has been estimated between 3-10% annually for RSV in older adults (Boyce et al., 2000) and 8-10% for influenza in adults (Tokars et al., 2018). Often, as is the case with influenza, older adults are at higher risk for hospitalization and death than other age groups (Czaja et al. 2019). There are comorbidities that are associated with increased hospitalization risk for older adults, such as congestive heart failure and chronic lung disease (Lee et al., 2013). Further, asthma, COPD, and congestive heart failure can exacerbate respiratory virus infections. Here we report counts for a selection of high-risk medical conditions such as chronic lung diseases and asthma.

Incidence rates in infants and children (defined here as individuals less than five years of age) with respiratory virus infections are higher than other age groups, except adults 65 and older (Centers for Disease Control and Prevention, 2023c; Centers for Disease Control and Prevention, 2023d). In the future, we plan to include high-risk comorbid states, such as congenital heart disease, preterm birth, and cystic fibrosis (Committee on Infectious Diseases and Bronchiolitis Guidelines Committee et al., 2014).

It is important for public health experts and clinical providers to understand the trends in these infections to inform decisions about public health, clinical care, and public policy. Connecting population-level trends with granular clinical information available in Truveta Studio can be very useful to more deeply understand which cohorts are most impacted.

This report is intended to supplement the surveillance data provided by the CDC (Centers for Disease Control and Prevention, 2023b). This report includes additional independent data and clinical detail that is not captured in other reports.

## Methods

### Data

We evaluated respiratory virus-associated hospitalization trends of six common respiratory viruses: COVID-19, influenza, human metapneumovirus (HMPV), parainfluenza virus, respiratory syncytial virus (RSV), and rhinovirus between October 01, 2019 and October 13, 2024 using a subset of Truveta Data. Truveta provides access to continuously updated, linked, and de-identified electronic health record (EHR) from a collective of US health care systems that provide 18% of all daily clinical care in the US, including structured information on demographics, encounters, diagnoses, vital signs (e.g., weight, BMI, blood pressure), medication requests (prescriptions), medication administration, laboratory and diagnostic tests and results (e.g., COVID tests and values), and procedures. Updated EHR data are provided daily to Truveta by constituent health care systems. The data used in this study was provided on October 24, 2024 and included de-identified patient care data primarily located across ten states: Virginia, New York, Florida, California, Illinois, Ohio, Washington, North Carolina, Wisconsin, and Louisiana.

### Population

We identified hospitalized patients who tested positive for one of the selected respiratory viruses within 14 days before or during the hospitalization. Positive lab results were identified using LOINC codes. Every respiratory virus-associated hospitalization has been grouped such that every hospitalization within 90 days is considered to be the same infection and thus only counted once. To align with seasonality in respiratory transmission, time periods include October 1st through September 30th of the following year.

### Hospitalization rate analysis

Characteristics of respiratory virus-associated hospitalized patients were summarized, including demographics and comorbidities by respiratory virus season. Characteristics are provided for the overall population, individual viruses, and two at-risk sub populations: infants and children (age 0-4) and older adults (age 65 and over).

Respiratory virus-associated hospitalizations rates were summarized over time by virus. The hospitalization rate was calculated weekly as the number of patients with a respiratory virus associated hospitalization divided by the number of patients with a hospital admission in that week. Patients were included in this calculation on the first day of their hospitalization. If their stay was greater than one day, they were not counted on subsequent dates.

Given the unadjusted nature of the data, the rates do not account for undertesting and other variability that exists across patient groups, providers, and systems. For further limitations, see the section below.

### Test positivity rate analysis

Lab results with known results were summarized over time by virus. The positivity rate was calculated monthly as the number of positive lab results divided by the total number of lab results with known results in that month.

Given the unadjusted nature of the data, the rates do not account for undertesting and other variability that exists across patient groups, providers, and systems. For further limitations, see the section below.

## Results

### Overall population

Our study population consists of 748,659 hospitalizations of 692,435 unique patients (Table 1).

**Table 1:** Patient characteristics by season.

|  | 2019/2020 | 2020/2021 | 2021/2022 | 2022/2023 | 2023/2024 | Overall |
| --- | --- | --- | --- | --- | --- | --- |
| N | 95,147<br>(100.00%) | 177,439<br>(100.00%) | 179,424<br>(100.00%) | 125,817<br>(100.00%) | 114,608<br>(100.00%) | 692,435<br>(100.00%) |
| Age Group |  |  |  |  |  |  |
| < 6 months | 1,710 (1.80%) | 1,303 (0.73%) | 2,493 (1.39%) | 3,120 (2.48%) | 2,404 (2.10%) | 11,030 (1.59%) |
| 6 - <12 months | 762 (0.80%) | 508 (0.29%) | 1,116 (0.62%) | 1,478 (1.17%) | 1,223 (1.07%) | 5,087 (0.73%) |
| 1 - <2 years | 968 (1.02%) | 867 (0.49%) | 1,693 (0.94%) | 2,042 (1.62%) | 1,703 (1.49%) | 7,273 (1.05%) |
| 2 - 4 years | 1,244 (1.31%) | 972 (0.55%) | 2,407 (1.34%) | 3,120 (2.48%) | 2,203 (1.92%) | 9,946 (1.44%) |
| 5 - 17 years | 1,398 (1.47%) | 1,777 (1.00%) | 3,082 (1.72%) | 3,532 (2.81%) | 3,027 (2.64%) | 12,816 (1.85%) |
| 18 - 49 years | 19,975 (20.99%) | 40,303 (22.71%) | 37,749 (21.04%) | 17,504 (13.91%) | 15,379 (13.42%) | 130,910<br>(18.91%) |
| 50 - 64 years | 23,099 (24.28%) | 44,973 (25.35%) | 36,897 (20.56%) | 19,935 (15.84%) | 18,802 (16.41%) | 143,706<br>(20.75%) |
| 65 - 74 years | 18,738 (19.69%) | 37,048 (20.88%) | 36,141 (20.14%) | 24,772 (19.69%) | 23,095 (20.15%) | 139,794<br>(20.19%) |
| 75 - 84 years | 16,684 (17.53%) | 31,558 (17.79%) | 35,108 (19.57%) | 29,195 (23.20%) | 27,570 (24.06%) | 140,115<br>(20.24%) |
| 85+ years | 10,569 (11.11%) | 18,130 (10.22%) | 22,738 (12.67%) | 21,119 (16.79%) | 19,202 (16.75%) | 91,758 (13.25%) |
| Sex |  |  |  |  |  |  |
| Female | 47,433 (49.85%) | 86,165 (48.56%) | 91,788 (51.16%) | 65,943 (52.41%) | 59,869 (52.24%) | 351,198<br>(50.72%) |
| Male | 46,667 (49.05%) | 89,597 (50.49%) | 85,986 (47.92%) | 58,917 (46.83%) | 53,605 (46.77%) | 334,772<br>(48.35%) |
| Other | 0 | 3 (0.00%) | 2 (0.00%) | 4 (0.00%) | 0 | 9 (0.00%) |
| Unknown | 1,047 (1.10%) | 1,674 (0.94%) | 1,648 (0.92%) | 953 (0.76%) | 1,134 (0.99%) | 6,456 (0.93%) |
| Race |  |  |  |  |  |  |
| White | 52,069 (54.72%) | 113,246<br>(63.82%) | 121,205<br>(67.55%) | 84,878 (67.46%) | 76,012 (66.32%) | 447,410<br>(64.61%) |
| Black or African American | 15,112 (15.88%) | 22,590 (12.73%) | 21,607 (12.04%) | 14,119 (11.22%) | 14,751 (12.87%) | 88,179 (12.73%) |
| Asian | 2,989 (3.14%) | 4,292 (2.42%) | 4,397 (2.45%) | 4,196 (3.34%) | 3,204 (2.80%) | 19,078 (2.76%) |
| Native Hawaiian or Other Pacific Islander | 590 (0.62%) | 843 (0.48%) | 936 (0.52%) | 680 (0.54%) | 563 (0.49%) | 3,612 (0.52%) |
| American Indian or Alaska Native | 613 (0.64%) | 1,056 (0.60%) | 1,125 (0.63%) | 833 (0.66%) | 670 (0.58%) | 4,297 (0.62%) |
| Other Race | 8,395 (8.82%) | 12,440 (7.01%) | 10,310 (5.75%) | 8,399 (6.68%) | 7,330 (6.40%) | 46,874 (6.77%) |
| Unknown | 15,379 (16.16%) | 22,972 (12.95%) | 19,844 (11.06%) | 12,712 (10.10%) | 12,078 (10.54%) | 82,985 (11.98%) |
| Ethnicity |  |  |  |  |  |  |
| Hispanic or Latino | 13,223 (13.90%) | 20,168 (11.37%) | 16,421 (9.15%) | 12,931 (10.28%) | 12,520 (10.92%) | 75,263 (10.87%) |
| Not Hispanic or Latino | 65,602 (68.95%) | 131,624 (74.18%) | 142,225 (79.27%) | 99,855 (79.37%) | 92,191 (80.44%) | 531,497 (76.76%) |
| Unknown | 16,322 (17.15%) | 25,647 (14.45%) | 20,778 (11.58%) | 13,031 (10.36%) | 9,897 (8.64%) | 85,675 (12.37%) |
| Virus |  |  |  |  |  |  |
| COVID | 61,944 (65.10%) | 166,877 (94.05%) | 156,018 (86.95%) | 76,040 (60.44%) | 61,610 (53.76%) | 522,489 (75.46%) |
| HMPV | 2,677 (2.81%) | 83 (0.05%) | 2,267 (1.26%) | 3,998 (3.18%) | 3,356 (2.93%) | 12,381 (1.79%) |
| RSV | 5,968 (6.27%) | 2,014 (1.14%) | 4,424 (2.47%) | 8,094 (6.43%) | 10,035 (8.76%) | 30,535 (4.41%) |
| influenza | 15,008 (15.77%) | 387 (0.22%) | 4,888 (2.72%) | 17,811 (14.16%) | 21,216 (18.51%) | 59,310 (8.57%) |
| parainfluenza virus | 1,402 (1.47%) | 1,456 (0.82%) | 2,404 (1.34%) | 4,388 (3.49%) | 3,757 (3.28%) | 13,407 (1.94%) |
| rhinovirus | 8,148 (8.56%) | 6,622 (3.73%) | 9,423 (5.25%) | 15,486 (12.31%) | 14,634 (12.77%) | 54,313 (7.84%) |
| Comorbidity |  |  |  |  |  |  |
| Asthma | 9,004 (9.46%) | 14,340 (8.08%) | 17,550 (9.78%) | 15,134 (12.03%) | 15,163 (13.23%) | 71,191 (10.28%) |
| Chronic Lung Disease | 6,419 (6.75%) | 11,057 (6.23%) | 12,099 (6.74%) | 9,592 (7.62%) | 9,508 (8.30%) | 48,675 (7.03%) |
| Chronic Obstructive Pulmonary Disease | 12,151 (12.77%) | 19,201 (10.82%) | 26,101 (14.55%) | 21,993 (17.48%) | 21,891 (19.10%) | 101,337 (14.63%) |

#### Hospitalization rate over time

The rate of respiratory virus-associated hospitalizations compared to all hospitalizations is shown in Figure 1. Figure 2 shows the same data stacked to represent the combined impact of the viruses.

**Figure 1:**
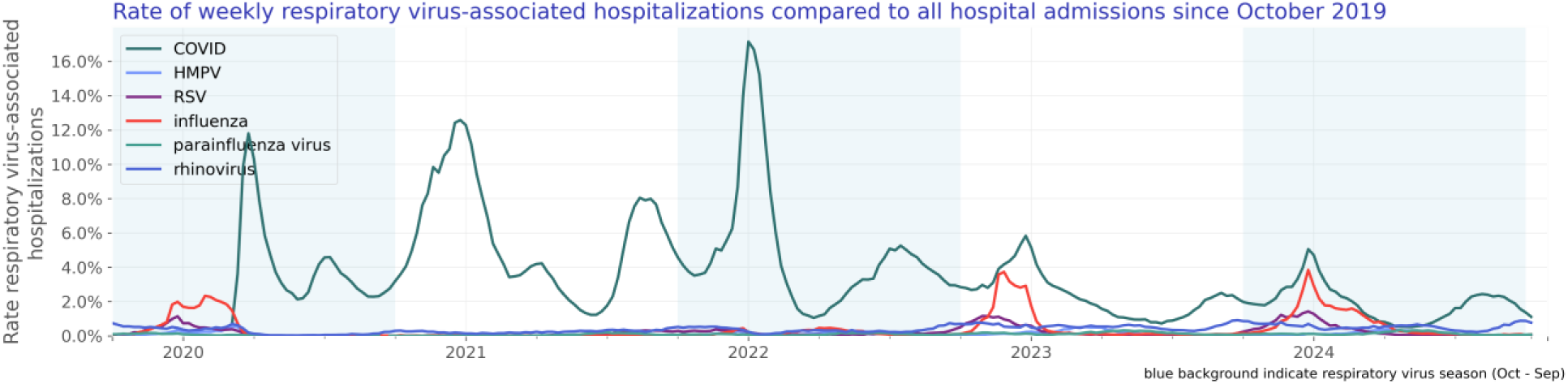
Rate of weekly respiratory virus-associated hospitalizations compared to all hospital admissions since October 2019

**Figure 2:**
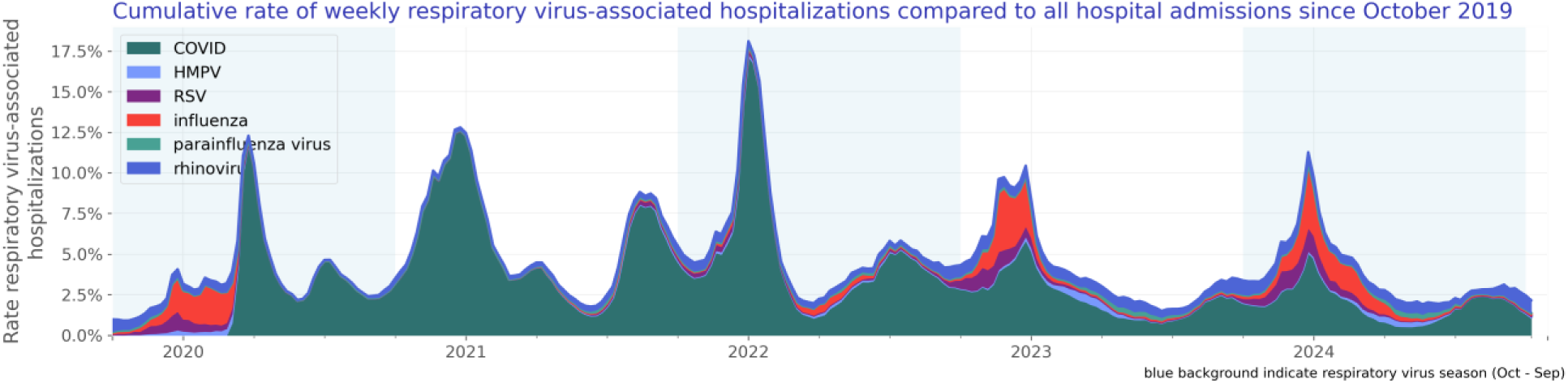
Cumulative rate of weekly respiratory virus-associated hospitalizations compared to all hospital admissions since October 2019

#### Test positivity rate over time

We included 52,947,476 lab results with known results of which 5,074,336 were positive. The test positivity rate is shown in Figure 3.

**Figure 3:**
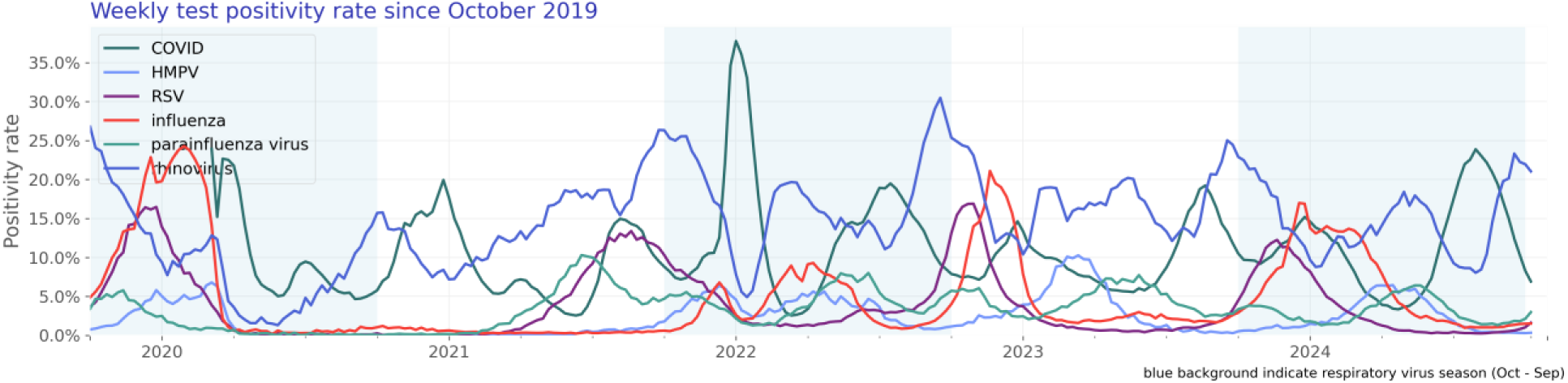
Weekly test positivity rate since October 2019

### COVID-19

Our COVID study population consists of 542,715 hospitalizations of 522,489 unique patients (Table 2).

**Table 2:** COVID patient characteristics by season.

|  | 2019/2020 | 2020/2021 | 2021/2022 | 2022/2023 | 2023/2024 | Overall |
| --- | --- | --- | --- | --- | --- | --- |
| N | 61,944<br>(100.00%) | 166,877<br>(100.00%) | 156,018<br>(100.00%) | 76,040<br>(100.00%) | 61,610<br>(100.00%) | 522,489<br>(100.00%) |
| Age Group |  |  |  |  |  |  |
| < 6 months | 115 (0.19%) | 327 (0.20%) | 785 (0.50%) | 433 (0.57%) | 364 (0.59%) | 2,024 (0.39%) |
| 6 - <12 months | 40 (0.06%) | 56 (0.03%) | 270 (0.17%) | 220 (0.29%) | 222 (0.36%) | 808 (0.15%) |
| 1 - <2 years | 58 (0.09%) | 101 (0.06%) | 277 (0.18%) | 196 (0.26%) | 259 (0.42%) | 891 (0.17%) |
| 2 - 4 years | 107 (0.17%) | 117 (0.07%) | 411 (0.26%) | 265 (0.35%) | 309 (0.50%) | 1,209 (0.23%) |
| 5 - 17 years | 365 (0.59%) | 916 (0.55%) | 1,393 (0.89%) | 583 (0.77%) | 632 (1.03%) | 3,889 (0.74%) |
| 18 - 49 years | 14,638 (23.63%) | 38,417 (23.02%) | 34,312 (21.99%) | 9,923 (13.05%) | 6,715 (10.90%) | 104,005<br>(19.91%) |
| 50 - 64 years | 16,177 (26.12%) | 43,344 (25.97%) | 33,422 (21.42%) | 11,564 (15.21%) | 8,940 (14.51%) | 113,447 (21.71%) |
| 65 - 74 years | 12,570 (20.29%) | 35,675 (21.38%) | 32,592 (20.89%) | 16,102 (21.18%) | 13,130 (21.31%) | 110,069 (21.07%) |
| 75 - 84 years | 11,078 (17.88%) | 30,452 (18.25%) | 31,888 (20.44%) | 20,847 (27.42%) | 17,876 (29.01%) | 112,141 (21.46%) |
| 85+ years | 6,796 (10.97%) | 17,472 (10.47%) | 20,668 (13.25%) | 15,907 (20.92%) | 13,163 (21.37%) | 74,006 (14.16%) |
| Sex |  |  |  |  |  |  |
| Female | 29,662 (47.89%) | 80,758 (48.39%) | 79,645 (51.05%) | 39,240 (51.60%) | 31,321 (50.84%) | 260,626 (49.88%) |
| Male | 31,605 (51.02%) | 84,526 (50.65%) | 74,902 (48.01%) | 36,188 (47.59%) | 29,595 (48.04%) | 256,816 (49.15%) |
| Other | 0 | 3 (0.00%) | 2 (0.00%) | 2 (0.00%) | 0 | 7 (0.00%) |
| Unknown | 677 (1.09%) | 1,590 (0.95%) | 1,469 (0.94%) | 610 (0.80%) | 694 (1.13%) | 5,040 (0.96%) |
| Race |  |  |  |  |  |  |
| White | 30,383 (49.05%) | 106,963 (64.10%) | 106,399 (68.20%) | 53,126 (69.87%) | 42,521 (69.02%) | 339,392 (64.96%) |
| Black or African American | 10,904 (17.60%) | 20,897 (12.52%) | 18,482 (11.85%) | 7,645 (10.05%) | 7,022 (11.40%) | 64,950 (12.43%) |
| Asian | 2,068 (3.34%) | 3,949 (2.37%) | 3,508 (2.25%) | 2,320 (3.05%) | 1,715 (2.78%) | 13,560 (2.60%) |
| Native Hawaiian or Other Pacific Islander | 406 (0.66%) | 774 (0.46%) | 737 (0.47%) | 311 (0.41%) | 212 (0.34%) | 2,440 (0.47%) |
| American Indian or Alaska Native | 397 (0.64%) | 985 (0.59%) | 942 (0.60%) | 415 (0.55%) | 336 (0.55%) | 3,075 (0.59%) |
| Other Race | 6,364 (10.27%) | 11,304 (6.77%) | 8,251 (5.29%) | 4,106 (5.40%) | 3,116 (5.06%) | 33,141 (6.34%) |
| Unknown | 11,422 (18.44%) | 22,005 (13.19%) | 17,699 (11.34%) | 8,117 (10.67%) | 6,688 (10.86%) | 65,931 (12.62%) |
| Ethnicity |  |  |  |  |  |  |
| Hispanic or Latino | 10,276 (16.59%) | 18,868 (11.31%) | 13,500 (8.65%) | 6,206 (8.16%) | 5,344 (8.67%) | 54,194 (10.37%) |
| Not Hispanic or Latino | 39,930 (64.46%) | 123,451 (73.98%) | 123,925 (79.43%) | 61,255 (80.56%) | 50,578 (82.09%) | 399,139 (76.39%) |
| Unknown | 11,738 (18.95%) | 24,558 (14.72%) | 18,593 (11.92%) | 8,579 (11.28%) | 5,688 (9.23%) | 69,156 (13.24%) |
| Comorbidity |  |  |  |  |  |  |
| Asthma | 4,081 (6.59%) | 12,697 (7.61%) | 14,001 (8.97%) | 7,494 (9.86%) | 6,884 (11.17%) | 45,157 (8.64%) |
| Chronic Lung Disease | 3,033 (4.90%) | 10,221 (6.12%) | 10,162 (6.51%) | 5,443 (7.16%) | 4,980 (8.08%) | 33,839 (6.48%) |
| Chronic Obstructive Pulmonary Disease | 5,213 (8.42%) | 17,539 (10.51%) | 22,096 (14.16%) | 12,873 (16.93%) | 11,734 (19.05%) | 69,455 (13.29%) |

#### Hospitalization rate over time

The rate of COVID-associated hospitalization is shown in Figure 4. Figure 5 shows seasonal trends.

**Figure 4:**
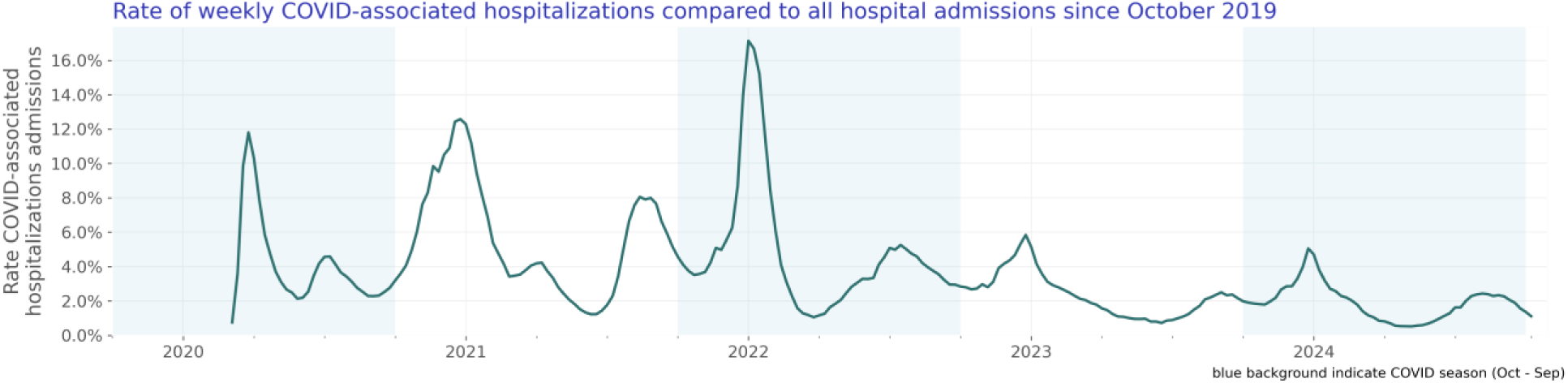
Rate of weekly COVID-associated hospitalizations compared to all hospital admissions since October 2019

**Figure 5:**
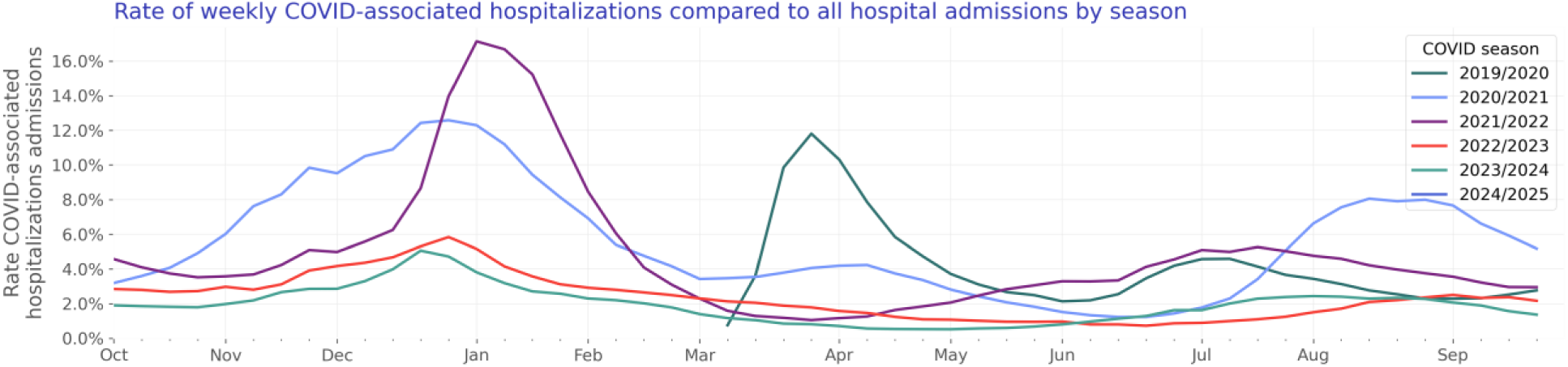
Rate of weekly COVID-associated hospitalizations compared to all hospital admissions by season

#### Test positivity rate over time

We included 30,120,112 COVID lab results with known results of which 3,497,812 were positive. The COVID test positivity rate is shown in Figure 6. Figure 7 shows yearly trends.

**Figure 6:**
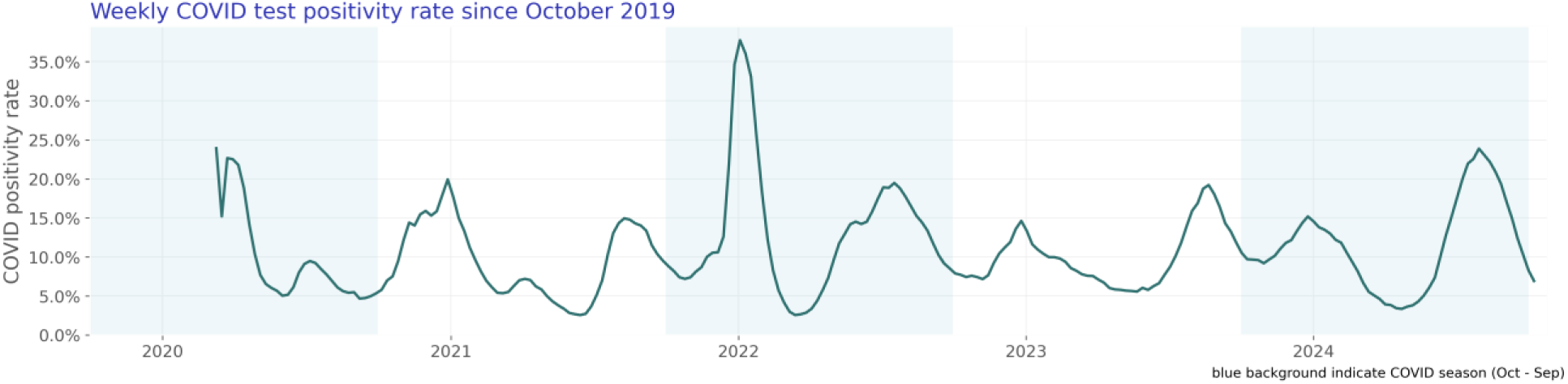
Weekly COVID test positivity rate since October 2019

**Figure 7:**
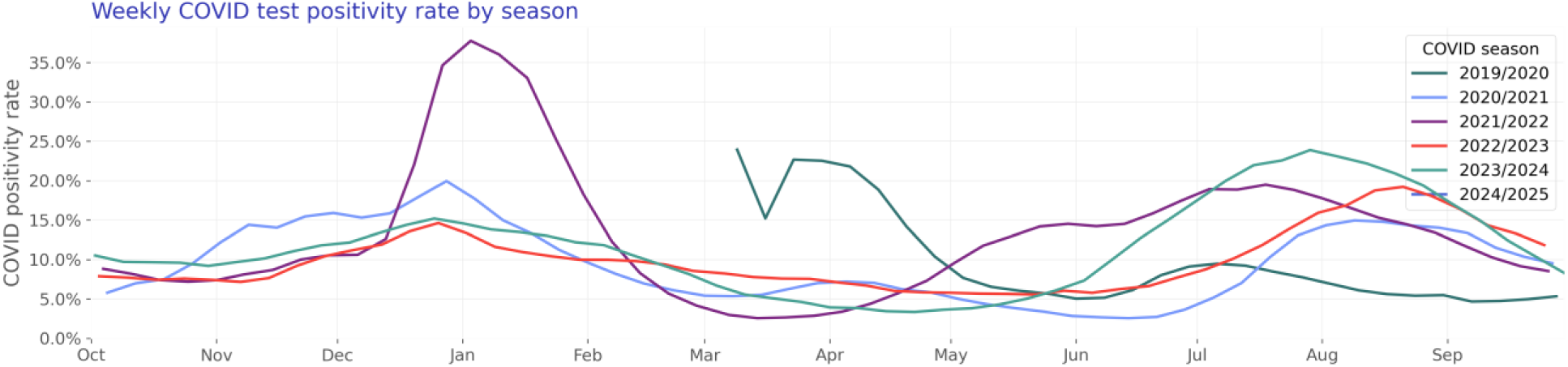
Weekly COVID test positivity rate by season

### Influenza

Our influenza study population consists of 68,774 hospitalizations of 59,310 unique patients (Table 3).

**Table 3:** Influenza patient characteristics by season.

|  | 2019/2020 | 2020/2021 | 2021/2022 | 2022/2023 | 2023/2024 | Overall |
| --- | --- | --- | --- | --- | --- | --- |
| N | 15,008<br>(100.00%) | 387 (100.00%) | 4,888 (100.00%) | 17,811<br>(100.00%) | 21,216<br>(100.00%) | 59,310<br>(100.00%) |
| Age Group |  |  |  |  |  |  |
| < 6 months | 86 (0.57%) | 1 (0.26%) | 27 (0.55%) | 75 (0.42%) | 85 (0.40%) | 274 (0.46%) |
| 6 - <12 months | 63 (0.42%) | 1 (0.26%) | 16 (0.33%) | 66 (0.37%) | 48 (0.23%) | 194 (0.33%) |
| 1 - <2 years | 104 (0.69%) | 0 | 29 (0.59%) | 120 (0.67%) | 84 (0.40%) | 337 (0.57%) |
| 2 - 4 years | 153 (1.02%) | 2 (0.52%) | 57 (1.17%) | 256 (1.44%) | 197 (0.93%) | 665 (1.12%) |
| 5 - 17 years | 329 (2.19%) | 3 (0.78%) | 173 (3.54%) | 583 (3.27%) | 510 (2.40%) | 1,598 (2.69%) |
| 18 - 49 years | 3,214 (21.42%) | 109 (28.17%) | 1,121 (22.93%) | 3,529 (19.81%) | 4,328 (20.40%) | 12,301 (20.74%) |
| 50 - 64 years | 3,821 (25.46%) | 99 (25.58%) | 904 (18.49%) | 3,741 (21.00%) | 4,812 (22.68%) | 13,377 (22.55%) |
| 65 - 74 years | 3,163 (21.08%) | 70 (18.09%) | 981 (20.07%) | 3,835 (21.53%) | 4,594 (21.65%) | 12,643 (21.32%) |
| 75 - 84 years | 2,545 (16.96%) | 73 (18.86%) | 954 (19.52%) | 3,581 (20.11%) | 4,255 (20.06%) | 11,408 (19.23%) |
| 85+ years | 1,530 (10.19%) | 29 (7.49%) | 626 (12.81%) | 2,025 (11.37%) | 2,303 (10.86%) | 6,513 (10.98%) |
| Sex |  |  |  |  |  |  |
| Female | 8,196 (54.61%) | 237 (61.24%) | 2,757 (56.40%) | 9,988 (56.08%) | 11,600 (54.68%) | 32,778 (55.27%) |
| Male | 6,637 (44.22%) | 145 (37.47%) | 2,089 (42.74%) | 7,711 (43.29%) | 9,452 (44.55%) | 26,034 (43.89%) |
| Other | 0 | 0 | 0 | 2 (0.01%) | 0 | 2 (0.00%) |
| Unknown | 175 (1.17%) | 5 (1.29%) | 42 (0.86%) | 110 (0.62%) | 164 (0.77%) | 496 (0.84%) |
| Race |  |  |  |  |  |  |
| White | 9,738 (64.89%) | 247 (63.82%) | 3,305 (67.61%) | 11,895 (66.78%) | 13,686 (64.51%) | 38,871 (65.54%) |
| Black or African American | 2,189 (14.59%) | 72 (18.60%) | 683 (13.97%) | 2,504 (14.06%) | 3,472 (16.37%) | 8,920 (15.04%) |
| Asian | 373 (2.49%) | 5 (1.29%) | 106 (2.17%) | 456 (2.56%) | 532 (2.51%) | 1,472 (2.48%) |
| Native Hawaiian or Other Pacific Islander | 74 (0.49%) | 1 (0.26%) | 37 (0.76%) | 120 (0.67%) | 145 (0.68%) | 377 (0.64%) |
| American Indian or Alaska Native | 82 (0.55%) | 2 (0.52%) | 45 (0.92%) | 157 (0.88%) | 132 (0.62%) | 418 (0.70%) |
| Other Race | 685 (4.56%) | 13 (3.36%) | 251 (5.14%) | 1,107 (6.22%) | 1,325 (6.25%) | 3,381 (5.70%) |
| Unknown | 1,867 (12.44%) | 47 (12.14%) | 461 (9.43%) | 1,572 (8.83%) | 1,924 (9.07%) | 5,871 (9.90%) |
| Ethnicity |  |  |  |  |  |  |
| Hispanic or Latino | 1,273 (8.48%) | 32 (8.27%) | 482 (9.86%) | 2,032 (11.41%) | 2,479 (11.68%) | 6,298 (10.62%) |
| Not Hispanic or Latino | 11,675 (77.79%) | 304 (78.55%) | 3,928 (80.36%) | 14,265 (80.09%) | 17,142 (80.80%) | 47,314 (79.77%) |
| Unknown | 2,060 (13.73%) | 51 (13.18%) | 478 (9.78%) | 1,514 (8.50%) | 1,595 (7.52%) | 5,698 (9.61%) |
| Comorbidity |  |  |  |  |  |  |
| Asthma | 2,038 (13.58%) | 35 (9.04%) | 735 (15.04%) | 2,700 (15.16%) | 3,253 (15.33%) | 8,761 (14.77%) |
| Chronic Lung Disease | 1,389 (9.26%) | 24 (6.20%) | 439 (8.98%) | 1,586 (8.90%) | 1,913 (9.02%) | 5,351 (9.02%) |
| Chronic Obstructive Pulmonary Disease | 3,046 (20.30%) | 61 (15.76%) | 1,033 (21.13%) | 3,777 (21.21%) | 4,381 (20.65%) | 12,298 (20.74%) |

#### Hospitalization rate over time

The rate of influenza-associated hospitalization is shown in Figure 8. Figure 9 shows seasonal trends.

**Figure 8:**
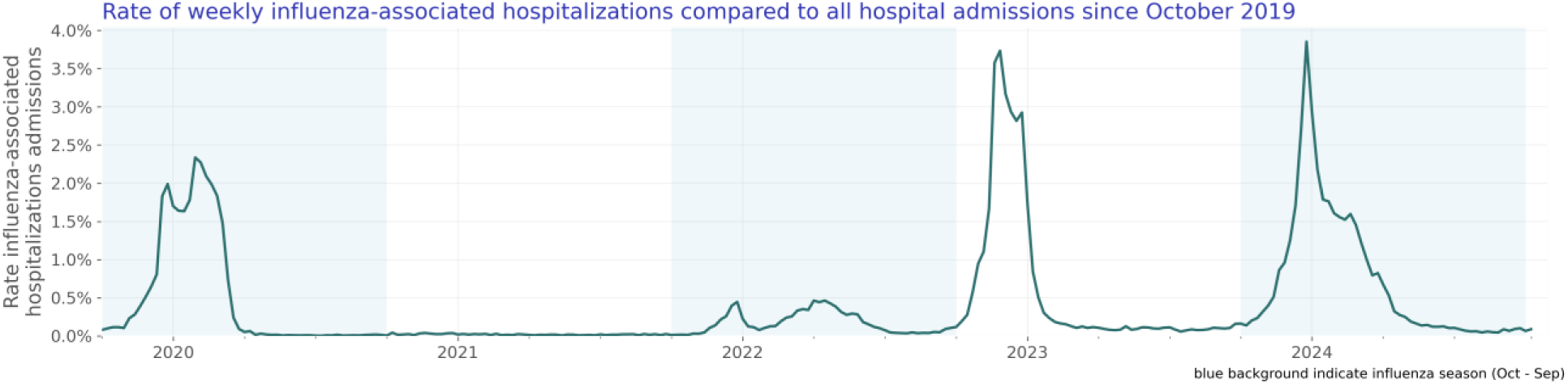
Rate of weekly influenza-associated hospitalizations compared to all hospital admissions since October 2019

**Figure 9:**
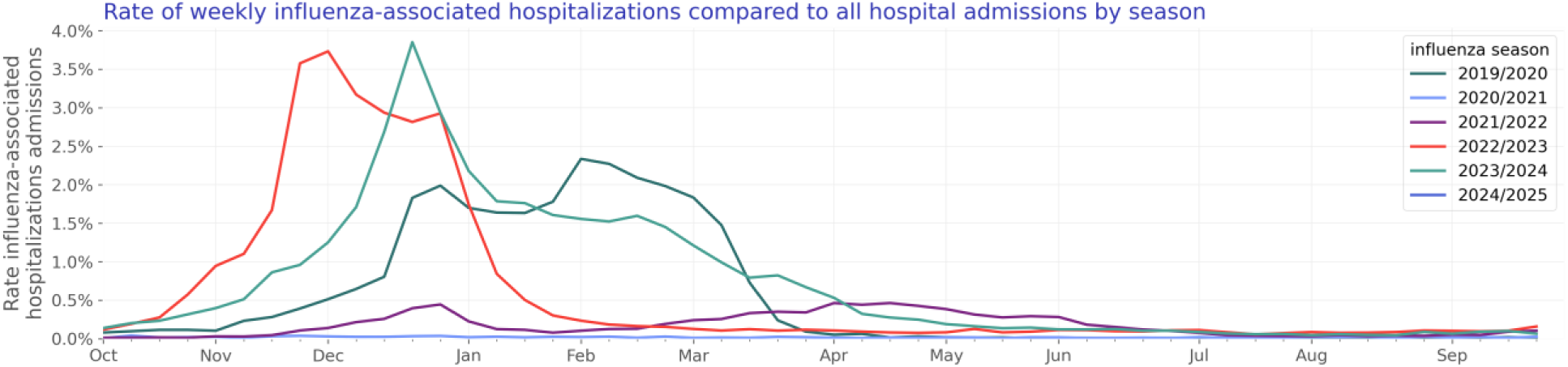
Rate of weekly influenza-associated hospitalizations compared to all hospital admissions by season

#### Test positivity rate over time

We included 12,752,701 influenza lab results with known results of which 997,708 were positive. The influenza test positivity rate is shown in Figure 10. Figure 11 shows yearly trends.

**Figure 10:**
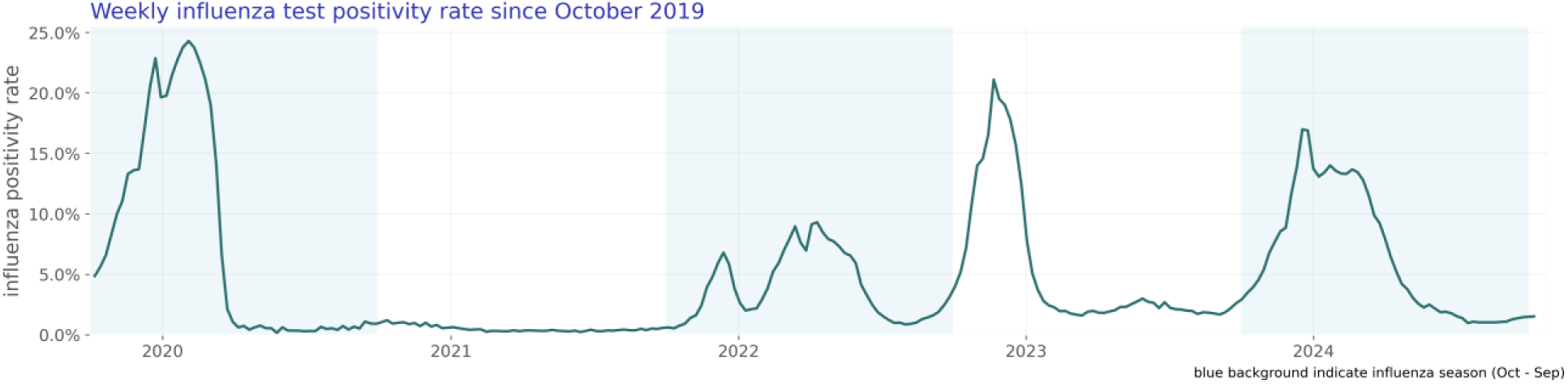
Weekly influenza test positivity rate since October 2019

**Figure 11:**
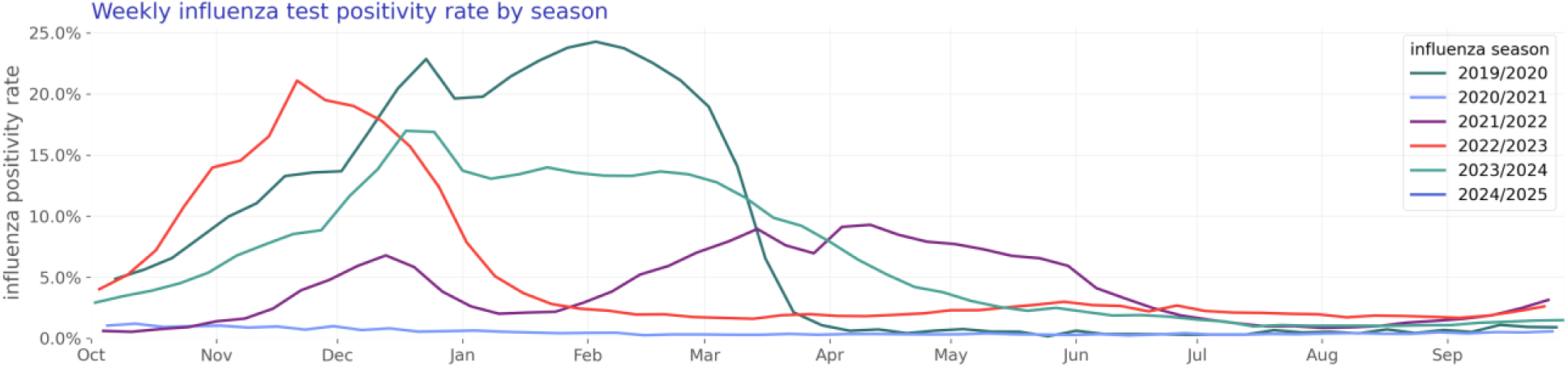
Weekly influenza test positivity rate by season

### Human metapneumovirus (HMPV)

Our HMPV study population consists of 16,925 hospitalizations of 12,381 unique patients (Table 4).

**Table 4:** HMPV patient characteristics by season.

|  | 2019/2020 | 2020/2021 | 2021/2022 | 2022/2023 | 2023/2024 | Overall |
| --- | --- | --- | --- | --- | --- | --- |
| N | 2,677 (100.00%) | 83 (100.00%) | 2,267 (100.00%) | 3,998 (100.00%) | 3,356 (100.00%) | 12,381 (100.00%) |
| Age Group |  |  |  |  |  |  |
| < 6 months | 47 (1.76%) | 2 (2.41%) | 56 (2.47%) | 134 (3.35%) | 69 (2.06%) | 308 (2.49%) |
| 6 - <12 months | 54 (2.02%) | 2 (2.41%) | 68 (3.00%) | 98 (2.45%) | 77 (2.29%) | 299 (2.41%) |
| 1 - <2 years | 64 (2.39%) | 7 (8.43%) | 119 (5.25%) | 139 (3.48%) | 79 (2.35%) | 408 (3.30%) |
| 2 - 4 years | 69 (2.58%) | 12 (14.46%) | 176 (7.76%) | 232 (5.80%) | 128 (3.81%) | 617 (4.98%) |
| 5 - 17 years | 51 (1.91%) | 0 | 93 (4.10%) | 130 (3.25%) | 91 (2.71%) | 365 (2.95%) |
| 18 - 49 years | 298 (11.13%) | 13 (15.66%) | 269 (11.87%) | 426 (10.66%) | 416 (12.40%) | 1,422 (11.49%) |
| 50 - 64 years | 556 (20.77%) | 19 (22.89%) | 419 (18.48%) | 681 (17.03%) | 595 (17.73%) | 2,270 (18.33%) |
| 65 - 74 years | 554 (20.69%) | 8 (9.64%) | 413 (18.22%) | 780 (19.51%) | 690 (20.56%) | 2,445 (19.75%) |
| 75 - 84 years | 580 (21.67%) | 14 (16.87%) | 418 (18.44%) | 844 (21.11%) | 745 (22.20%) | 2,601 (21.01%) |
| 85+ years | 404 (15.09%) | 6 (7.23%) | 236 (10.41%) | 534 (13.36%) | 466 (13.89%) | 1,646 (13.29%) |
| Sex |  |  |  |  |  |  |
| Female | 1,541 (57.56%) | 41 (49.40%) | 1,302 (57.43%) | 2,323 (58.10%) | 2,048 (61.03%) | 7,255 (58.60%) |
| Male | 1,099 (41.05%) | 40 (48.19%) | 951 (41.95%) | 1,652 (41.32%) | 1,285 (38.29%) | 5,027 (40.60%) |
| Unknown | 37 (1.38%) | 2 (2.41%) | 14 (0.62%) | 23 (0.58%) | 23 (0.69%) | 99 (0.80%) |
| Race |  |  |  |  |  |  |
| White | 2,005 (74.90%) | 56 (67.47%) | 1,647 (72.65%) | 2,861 (71.56%) | 2,236 (66.63%) | 8,805 (71.12%) |
| Black or African American | 223 (8.33%) | 8 (9.64%) | 245 (10.81%) | 357 (8.93%) | 402 (11.98%) | 1,235 (9.97%) |
| Asian | 49 (1.83%) | 0 | 47 (2.07%) | 114 (2.85%) | 73 (2.18%) | 283 (2.29%) |
| Native Hawaiian or Other Pacific Islander | 13 (0.49%) | 0 | 15 (0.66%) | 14 (0.35%) | 8 (0.24%) | 50 (0.40%) |
| American Indian or Alaska Native | 17 (0.64%) | 0 | 12 (0.53%) | 36 (0.90%) | 18 (0.54%) | 83 (0.67%) |
| Other Race | 105 (3.92%) | 9 (10.84%) | 111 (4.90%) | 290 (7.25%) | 262 (7.81%) | 777 (6.28%) |
| Unknown | 265 (9.90%) | 10 (12.05%) | 190 (8.38%) | 326 (8.15%) | 357 (10.64%) | 1,148 (9.27%) |
| Ethnicity |  |  |  |  |  |  |
| Hispanic or Latino | 180 (6.72%) | 17 (20.48%) | 221 (9.75%) | 536 (13.41%) | 485 (14.45%) | 1,439 (11.62%) |
| Not Hispanic or Latino | 2,162 (80.76%) | 53 (63.86%) | 1,829 (80.68%) | 3,153 (78.86%) | 2,626 (78.25%) | 9,823 (79.34%) |
| Unknown | 335 (12.51%) | 13 (15.66%) | 217 (9.57%) | 309 (7.73%) | 245 (7.30%) | 1,119 (9.04%) |
| Comorbidity |  |  |  |  |  |  |
| Asthma | 429 (16.03%) | 10 (12.05%) | 348 (15.35%) | 631 (15.78%) | 534 (15.91%) | 1,952 (15.77%) |
| Chronic Lung Disease | 364 (13.60%) | 10 (12.05%) | 272 (12.00%) | 418 (10.46%) | 315 (9.39%) | 1,379 (11.14%) |
| Chronic Obstructive Pulmonary Disease | 637 (23.80%) | 13 (15.66%) | 481 (21.22%) | 801 (20.04%) | 643 (19.16%) | 2,575 (20.80%) |

#### Hospitalization rate over time

The rate of HMPV-associated hospitalization is shown in Figure 12. Figure 13 shows seasonal trends.

**Figure 12:**
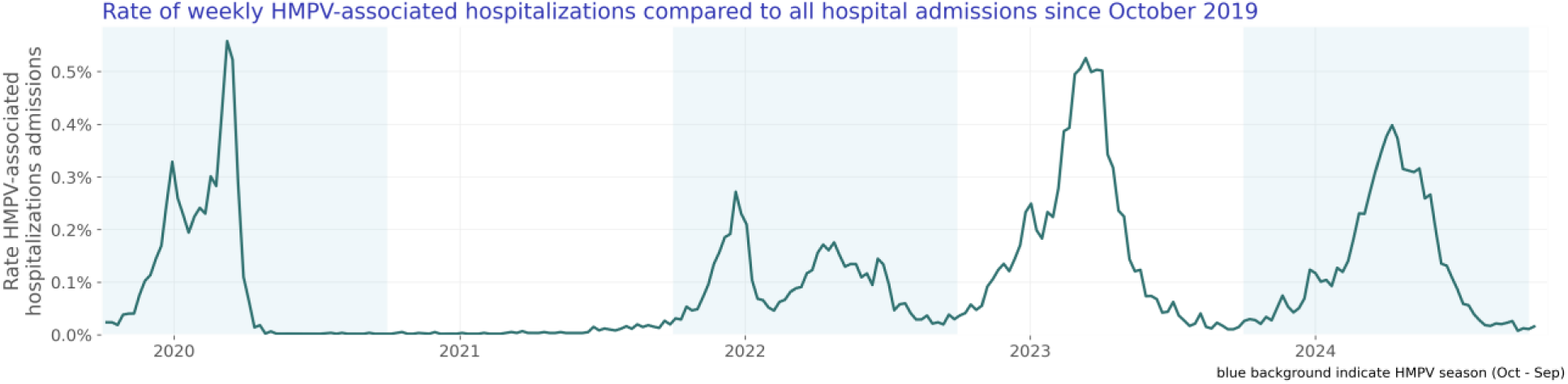
Rate of weekly HMPV-associated hospitalizations compared to all hospital admissions since October 2019

**Figure 13:**
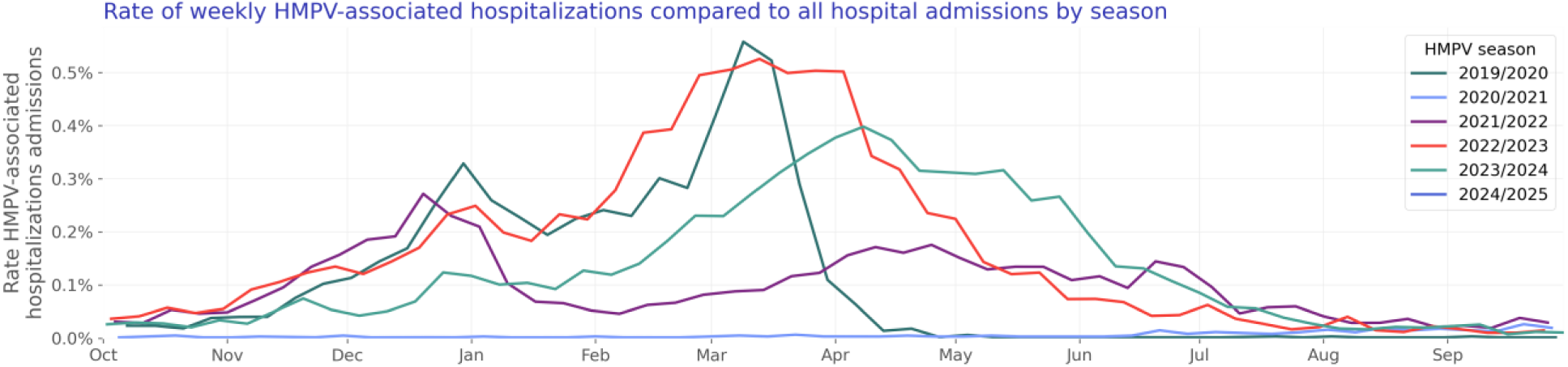
Rate of weekly HMPV-associated hospitalizations compared to all hospital admissions by season

#### Test positivity rate over time

We included 1,604,618 HMPV lab results with known results of which 43,099 were positive. The HMPV test positivity rate is shown in Figure 14. Figure 15 shows yearly trends.

**Figure 14:**
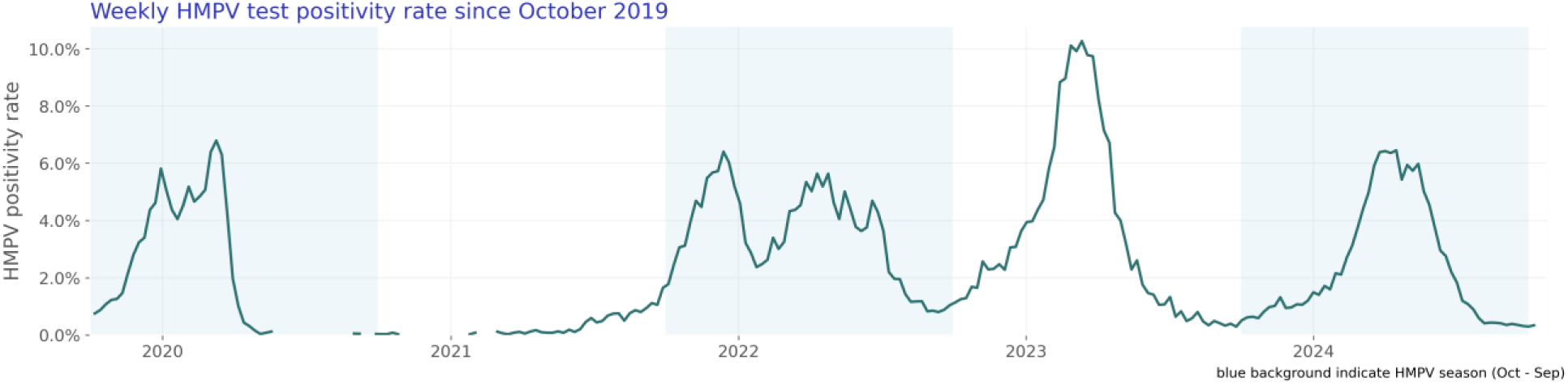
Weekly HMPV test positivity rate since October 2019

**Figure 15:**
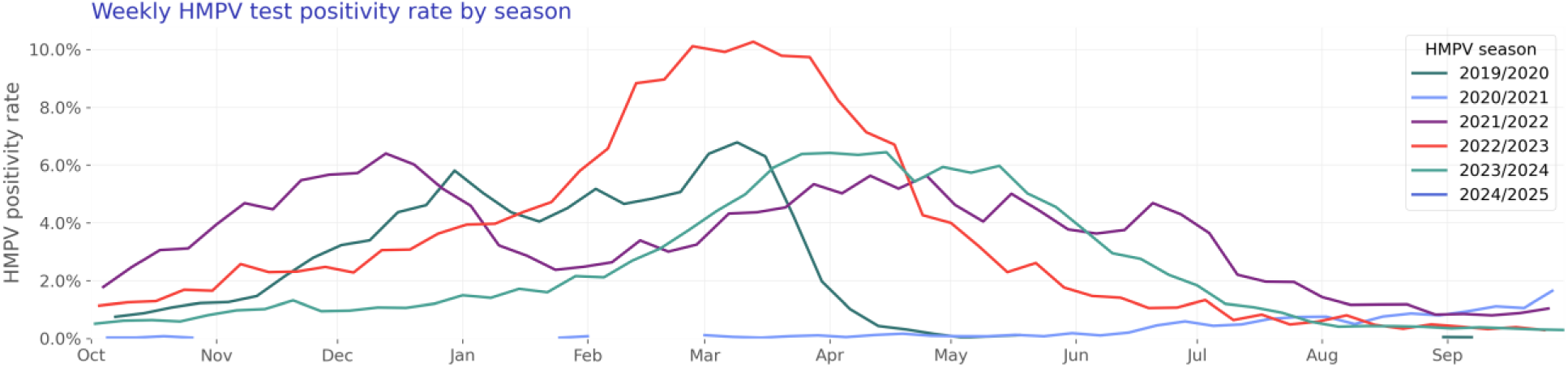
Weekly HMPV test positivity rate by season

#### Parainfluenza virus

Our parainfluenza virus study population consists of 16,315 hospitalizations of 13,407 unique patients (Table 5).

**Table 5:** Parainfluenza virus patient characteristics by season.

|  | 2019/2020 | 2020/2021 | 2021/2022 | 2022/2023 | 2023/2024 | Overall |
| --- | --- | --- | --- | --- | --- | --- |
| N | 1,402 (100.00%) | 1,456 (100.00%) | 2,404 (100.00%) | 4,388 (100.00%) | 3,757 (100.00%) | 13,407 (100.00%) |
| Age Group |  |  |  |  |  |  |
| < 6 months | 74 (5.28%) | 81 (5.56%) | 123 (5.12%) | 209 (4.76%) | 121 (3.22%) | 608 (4.53%) |
| 6 - <12 months | 33 (2.35%) | 64 (4.40%) | 90 (3.74%) | 145 (3.30%) | 79 (2.10%) | 411 (3.07%) |
| 1 - <2 years | 54 (3.85%) | 94 (6.46%) | 118 (4.91%) | 170 (3.87%) | 129 (3.43%) | 565 (4.21%) |
| 2 - 4 years | 79 (5.63%) | 109 (7.49%) | 194 (8.07%) | 257 (5.86%) | 151 (4.02%) | 790 (5.89%) |
| 5 - 17 years | 62 (4.42%) | 84 (5.77%) | 126 (5.24%) | 212 (4.83%) | 125 (3.33%) | 609 (4.54%) |
| 18 - 49 years | 144 (10.27%) | 193 (13.26%) | 297 (12.35%) | 506 (11.53%) | 413 (10.99%) | 1,553 (11.58%) |
| 50 - 64 years | 264 (18.83%) | 268 (18.41%) | 421 (17.51%) | 741 (16.89%) | 691 (18.39%) | 2,385 (17.79%) |
| 65 - 74 years | 245 (17.48%) | 239 (16.41%) | 427 (17.76%) | 763 (17.39%) | 742 (19.75%) | 2,416 (18.02%) |
| 75 - 84 years | 255 (18.19%) | 210 (14.42%) | 356 (14.81%) | 822 (18.73%) | 812 (21.61%) | 2,455 (18.31%) |
| 85+ years | 192 (13.69%) | 114 (7.83%) | 252 (10.48%) | 563 (12.83%) | 494 (13.15%) | 1,615 (12.05%) |
| Sex |  |  |  |  |  |  |
| Female | 761 (54.28%) | 753 (51.72%) | 1,313 (54.62%) | 2,358 (53.74%) | 2,040 (54.30%) | 7,225 (53.89%) |
| Male | 626 (44.65%) | 690 (47.39%) | 1,072 (44.59%) | 1,993 (45.42%) | 1,677 (44.64%) | 6,058 (45.19%) |
| Unknown | 15 (1.07%) | 13 (0.89%) | 19 (0.79%) | 37 (0.84%) | 40 (1.06%) | 124 (0.92%) |
| Race |  |  |  |  |  |  |
| White | 935 (66.69%) | 910 (62.50%) | 1,511 (62.85%) | 2,783 (63.42%) | 2,361 (62.84%) | 8,500 (63.40%) |
| Black or African American | 135 (9.63%) | 200 (13.74%) | 290 (12.06%) | 492 (11.21%) | 484 (12.88%) | 1,601 (11.94%) |
| Asian | 55 (3.92%) | 40 (2.75%) | 100 (4.16%) | 191 (4.35%) | 106 (2.82%) | 492 (3.67%) |
| Native Hawaiian or Other Pacific Islander | 14 (1.00%) | 4 (0.27%) | 11 (0.46%) | 21 (0.48%) | 27 (0.72%) | 77 (0.57%) |
| American Indian or Alaska Native | 9 (0.64%) | 14 (0.96%) | 20 (0.83%) | 37 (0.84%) | 26 (0.69%) | 106 (0.79%) |
| Other Race | 87 (6.21%) | 142 (9.75%) | 226 (9.40%) | 440 (10.03%) | 340 (9.05%) | 1,235 (9.21%) |
| Unknown | 167 (11.91%) | 146 (10.03%) | 246 (10.23%) | 424 (9.66%) | 413 (10.99%) | 1,396 (10.41%) |
| Ethnicity |  |  |  |  |  |  |
| Hispanic or Latino | 102 (7.28%) | 157 (10.78%) | 302 (12.56%) | 563 (12.83%) | 543 (14.45%) | 1,667 (12.43%) |
| Not Hispanic or Latino | 1,110 (79.17%) | 1,132 (77.75%) | 1,849 (76.91%) | 3,403 (77.55%) | 2,935 (78.12%) | 10,429 (77.79%) |
| Unknown | 190 (13.55%) | 167 (11.47%) | 253 (10.52%) | 422 (9.62%) | 279 (7.43%) | 1,311 (9.78%) |
| Comorbidity |  |  |  |  |  |  |
| Asthma | 225 (16.05%) | 222 (15.25%) | 374 (15.56%) | 682 (15.54%) | 586 (15.60%) | 2,089 (15.58%) |
| Chronic Lung Disease | 178 (12.70%) | 137 (9.41%) | 262 (10.90%) | 425 (9.69%) | 413 (10.99%) | 1,415 (10.55%) |
| Chronic Obstructive Pulmonary Disease | 322 (22.97%) | 304 (20.88%) | 505 (21.01%) | 903 (20.58%) | 875 (23.29%) | 2,909 (21.70%) |

#### Hospitalization rate over time

The rate of parainfluenza virus-associated hospitalization is shown in Figure 16. Figure 17 shows seasonal trends.

**Figure 16:**
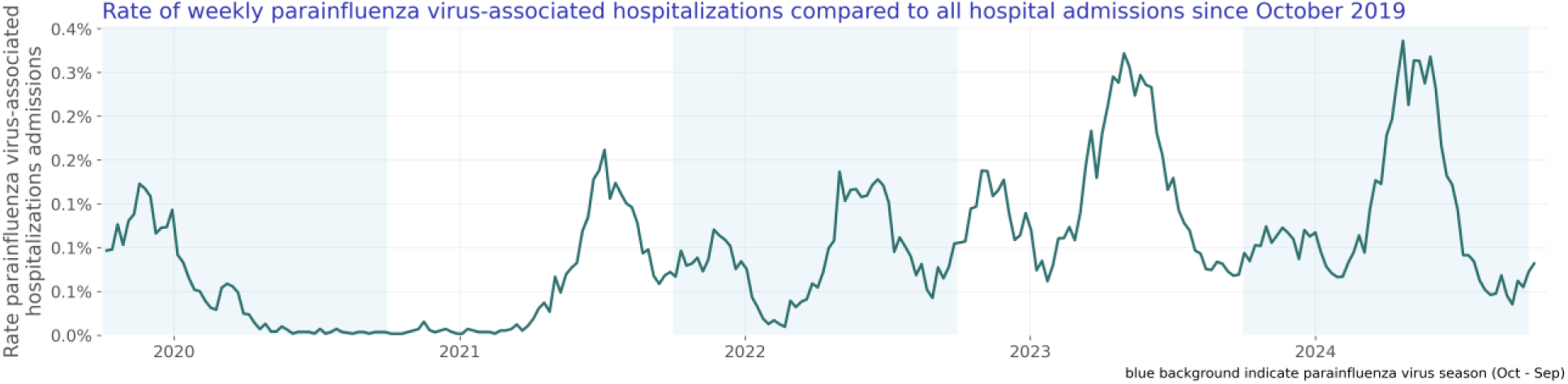
Rate of weekly parainfluenza virus-associated hospitalizations compared to all hospital admissions since October 2019

**Figure 17:**
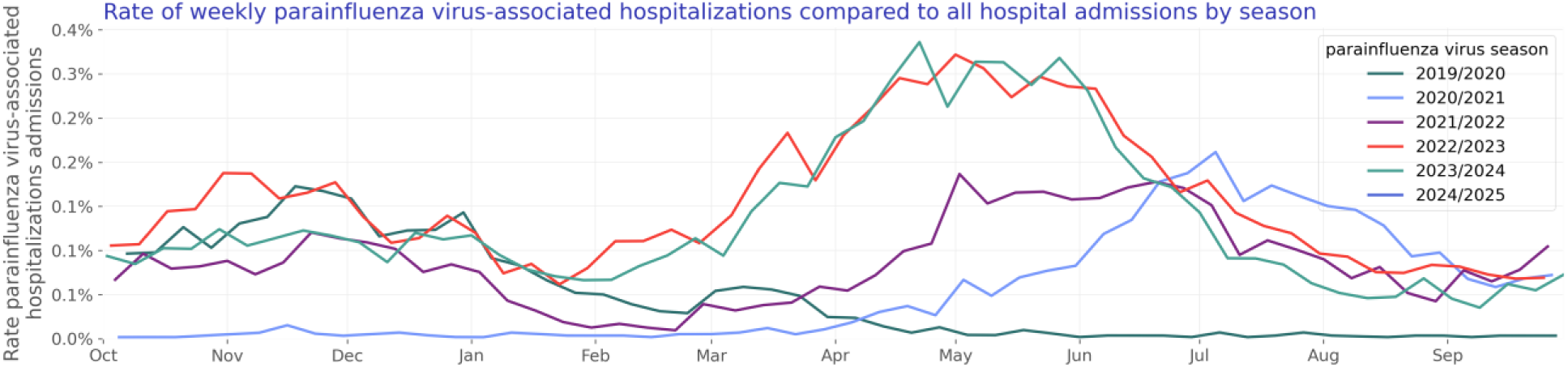
Rate of weekly parainfluenza virus-associated hospitalizations compared to all hospital admissions by season

#### Test positivity rate over time

We included 1,687,544 parainfluenza virus lab results with known results of which 60,220 were positive. The parainfluenza virus test positivity rate is shown in Figure 18. Figure 19 shows yearly trends.

**Figure 18:**
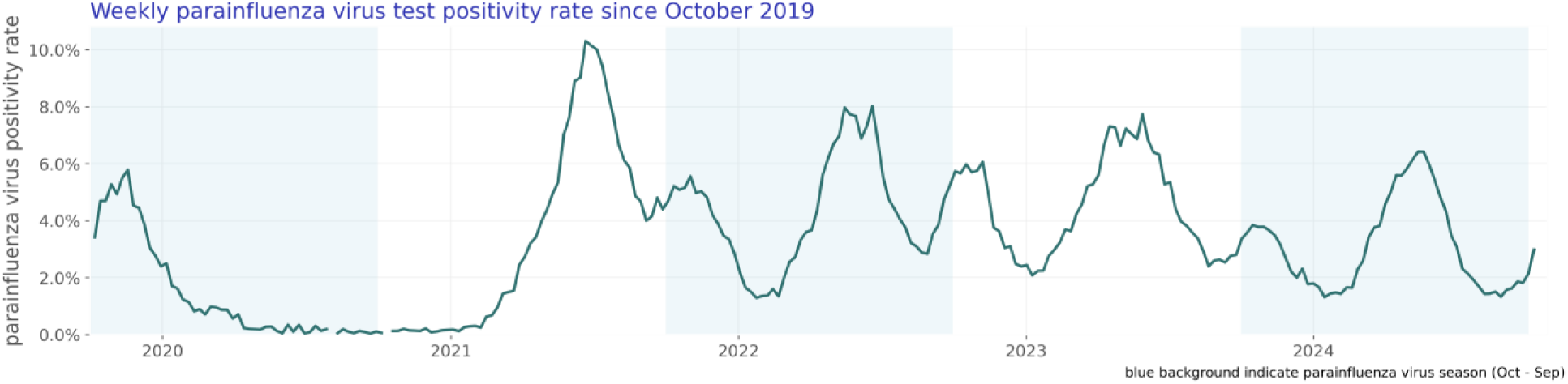
Weekly parainfluenza virus test positivity rate since October 2019

**Figure 19:**
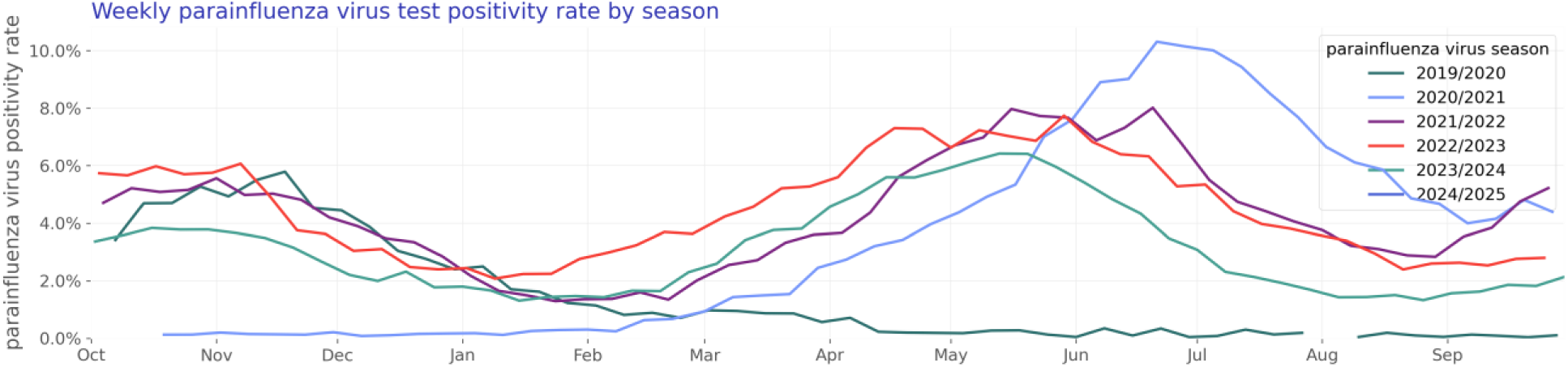
Weekly parainfluenza virus test positivity rate by season

### Respiratory syncytial virus (RSV)

Our RSV study population consists of 37,638 hospitalizations of 30,535 unique patients (Table 6).

**Table 6:** RSV patient characteristics by season.

|  | 2019/2020 | 2020/2021 | 2021/2022 | 2022/2023 | 2023/2024 | Overall |
| --- | --- | --- | --- | --- | --- | --- |
| N | 5,968 (100.00%) | 2,014 (100.00%) | 4,424 (100.00%) | 8,094 (100.00%) | 10,035 (100.00%) | 30,535 (100.00%) |
| Age Group |  |  |  |  |  |  |
| < 6 months | 962 (16.12%) | 507 (25.17%) | 801 (18.11%) | 1,407 (17.38%) | 1,094 (10.90%) | 4,771 (15.62%) |
| 6 - <12 months | 353 (5.91%) | 138 (6.85%) | 288 (6.51%) | 510 (6.30%) | 427 (4.26%) | 1,716 (5.62%) |
| 1 - <2 years | 341 (5.71%) | 182 (9.04%) | 359 (8.11%) | 569 (7.03%) | 463 (4.61%) | 1,914 (6.27%) |
| 2 - 4 years | 346 (5.80%) | 151 (7.50%) | 363 (8.21%) | 714 (8.82%) | 418 (4.17%) | 1,992 (6.52%) |
| 5 - 17 years | 101 (1.69%) | 47 (2.33%) | 123 (2.78%) | 260 (3.21%) | 217 (2.16%) | 748 (2.45%) |
| 18 - 49 years | 341 (5.71%) | 186 (9.24%) | 387 (8.75%) | 617 (7.62%) | 889 (8.86%) | 2,420 (7.93%) |
| 50 - 64 years | 747 (12.52%) | 258 (12.81%) | 546 (12.34%) | 939 (11.60%) | 1,490 (14.85%) | 3,980 (13.03%) |
| 65 - 74 years | 900 (15.08%) | 238 (11.82%) | 575 (13.00%) | 1,094 (13.52%) | 1,677 (16.71%) | 4,484 (14.68%) |
| 75 - 84 years | 1,063 (17.81%) | 190 (9.43%) | 593 (13.40%) | 1,158 (14.31%) | 1,921 (19.14%) | 4,925 (16.13%) |
| 85+ years | 814 (13.64%) | 117 (5.81%) | 389 (8.79%) | 826 (10.21%) | 1,439 (14.34%) | 3,585 (11.74%) |
| Sex |  |  |  |  |  |  |
| Female | 3,265 (54.71%) | 1,078 (53.53%) | 2,280 (51.54%) | 4,331 (53.51%) | 5,526 (55.07%) | 16,480 (53.97%) |
| Male | 2,667 (44.69%) | 929 (46.13%) | 2,116 (47.83%) | 3,704 (45.76%) | 4,428 (44.13%) | 13,844 (45.34%) |
| Unknown | 36 (0.60%) | 7 (0.35%) | 28 (0.63%) | 59 (0.73%) | 81 (0.81%) | 211 (0.69%) |
| Race |  |  |  |  |  |  |
| White | 3,871 (64.86%) | 1,196 (59.38%) | 2,865 (64.76%) | 5,199 (64.23%) | 6,656 (66.33%) | 19,787 (64.80%) |
| Black or African American | 631 (10.57%) | 364 (18.07%) | 521 (11.78%) | 889 (10.98%) | 1,165 (11.61%) | 3,570 (11.69%) |
| Asian | 181 (3.03%) | 53 (2.63%) | 197 (4.45%) | 442 (5.46%) | 358 (3.57%) | 1,231 (4.03%) |
| Native Hawaiian or Other Pacific Islander | 30 (0.50%) | 17 (0.84%) | 72 (1.63%) | 145 (1.79%) | 103 (1.03%) | 367 (1.20%) |
| American Indian or Alaska Native | 47 (0.79%) | 14 (0.70%) | 40 (0.90%) | 63 (0.78%) | 78 (0.78%) | 242 (0.79%) |
| Other Race | 507 (8.50%) | 179 (8.89%) | 331 (7.48%) | 658 (8.13%) | 642 (6.40%) | 2,317 (7.59%) |
| Unknown | 701 (11.75%) | 191 (9.48%) | 398 (9.00%) | 698 (8.62%) | 1,033 (10.29%) | 3,021 (9.89%) |
| Ethnicity |  |  |  |  |  |  |
| Hispanic or Latino | 657 (11.01%) | 269 (13.36%) | 586 (13.25%) | 1,194 (14.75%) | 1,242 (12.38%) | 3,948 (12.93%) |
| Not Hispanic or Latino | 4,438 (74.36%) | 1,529 (75.92%) | 3,452 (78.03%) | 6,183 (76.39%) | 7,915 (78.87%) | 23,517 (77.02%) |
| Unknown | 873 (14.63%) | 216 (10.72%) | 386 (8.73%) | 717 (8.86%) | 878 (8.75%) | 3,070 (10.05%) |
| Comorbidity |  |  |  |  |  |  |
| Asthma | 791 (13.25%) | 224 (11.12%) | 565 (12.77%) | 1,047 (12.94%) | 1,419 (14.14%) | 4,046 (13.25%) |
| Chronic Lung Disease | 499 (8.36%) | 125 (6.21%) | 295 (6.67%) | 577 (7.13%) | 820 (8.17%) | 2,316 (7.58%) |
| Chronic Obstructive Pulmonary Disease | 1,193 (19.99%) | 263 (13.06%) | 720 (16.27%) | 1,361 (16.81%) | 1,998 (19.91%) | 5,535 (18.13%) |

#### Hospitalization rate over time

The rate of RSV-associated hospitalization is shown in Figure 20. Figure 21 shows seasonal trends.

**Figure 20:**
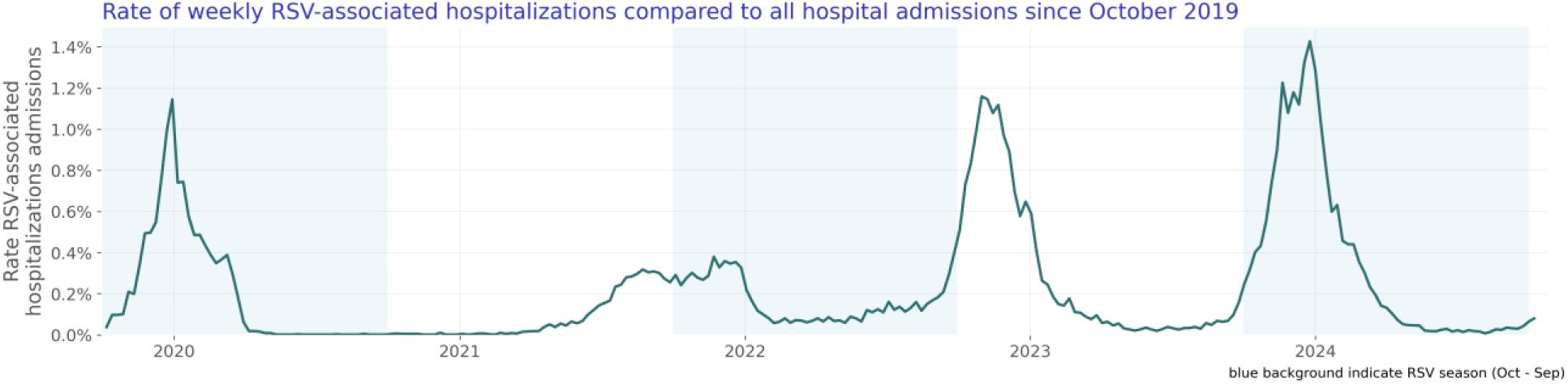
Rate of weekly RSV-associated hospitalizations compared to all hospital admissions since October 2019

**Figure 21:**
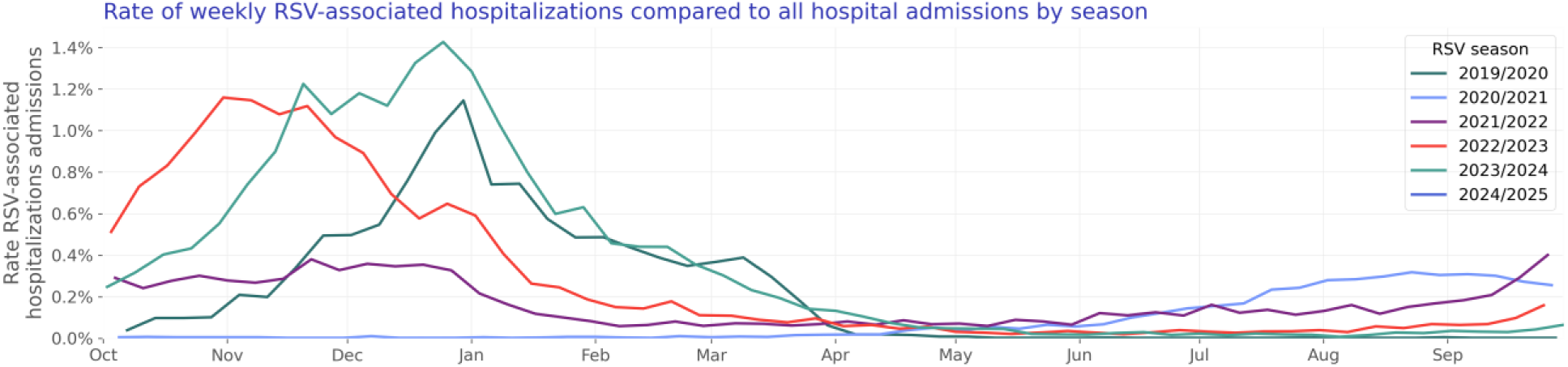
Rate of weekly RSV-associated hospitalizations compared to all hospital admissions by season

#### Test positivity rate over time

We included 5,243,725 RSV lab results with known results of which 239,431 were positive. The RSV test positivity rate is shown in Figure 22. Figure 23 shows yearly trends.

**Figure 22:**
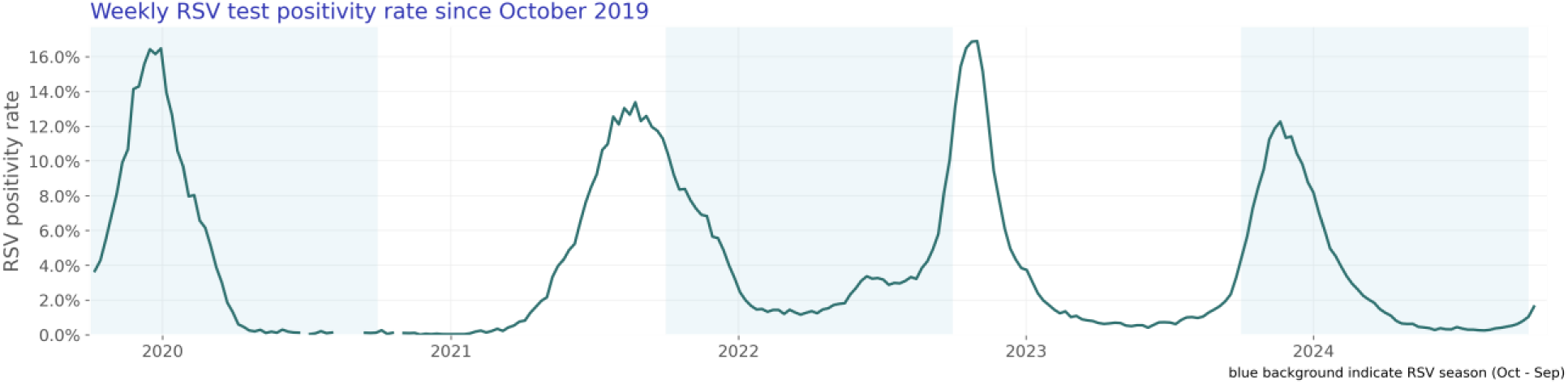
Weekly RSV test positivity rate since October 2019

**Figure 23:**
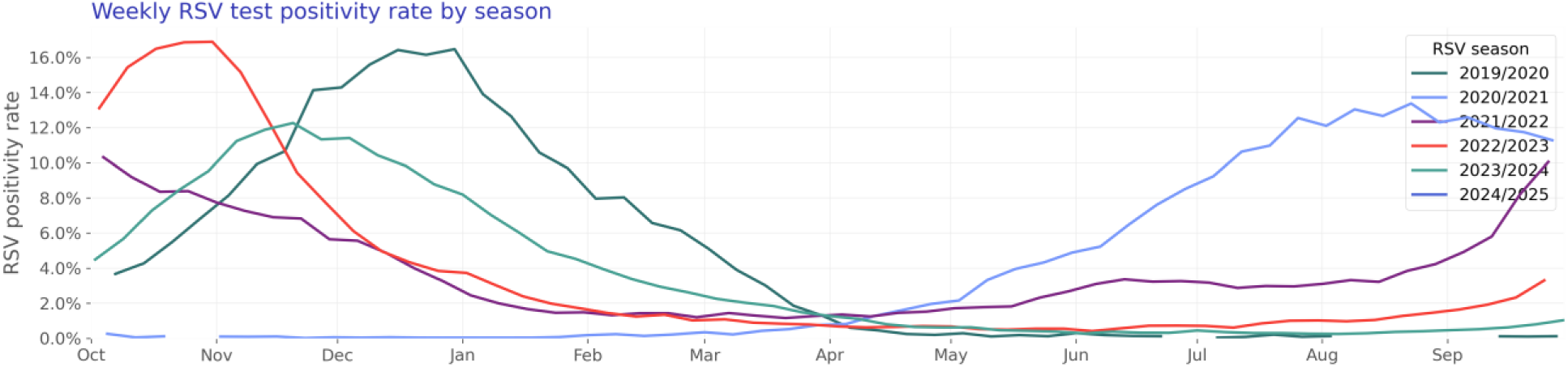
Weekly RSV test positivity rate by season

### Rhinovirus

Our rhinovirus study population consists of 66,292 hospitalizations of 54,313 unique patients (Table 7).

**Table 7:**
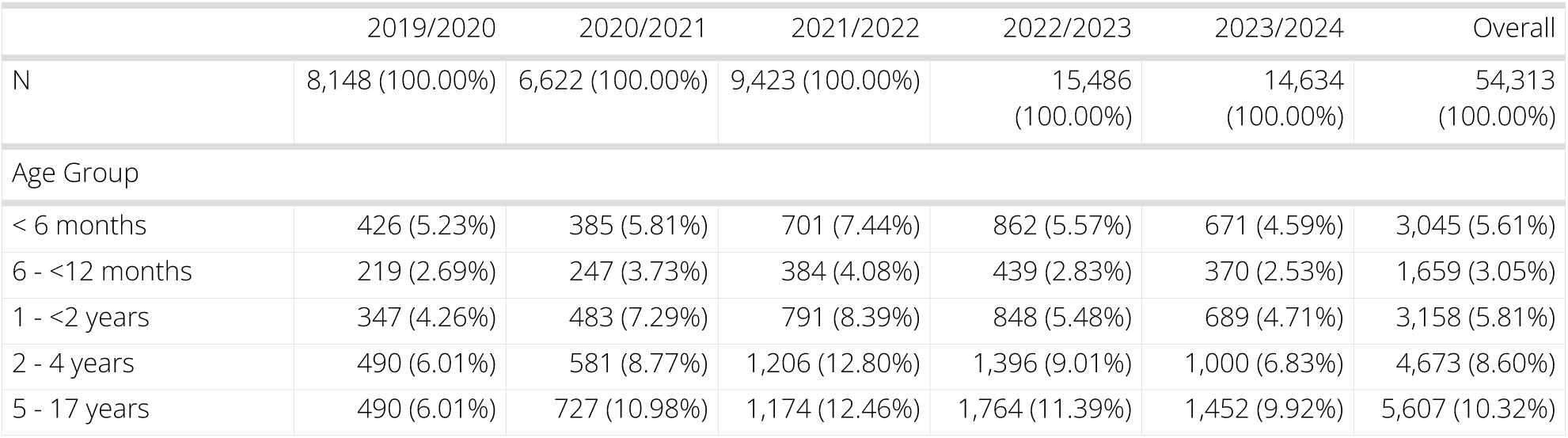
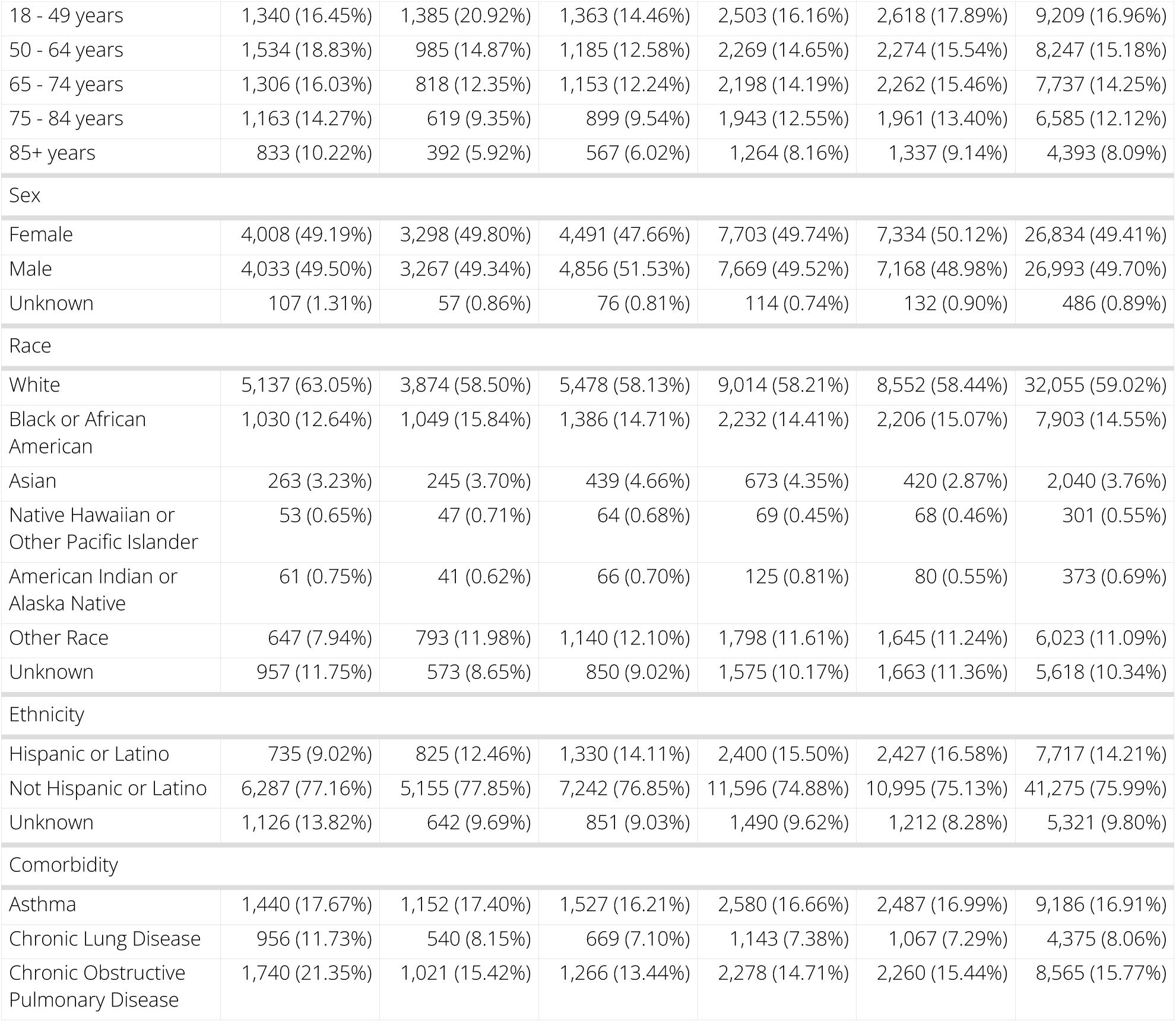
Rhinovirus patient characteristics by season.

#### Hospitalization rate over time

*The rate of rhinovirus-associated hospitalization is shown in Figure 24. Figure 25 shows seasonal trends.*

**Figure 24:**
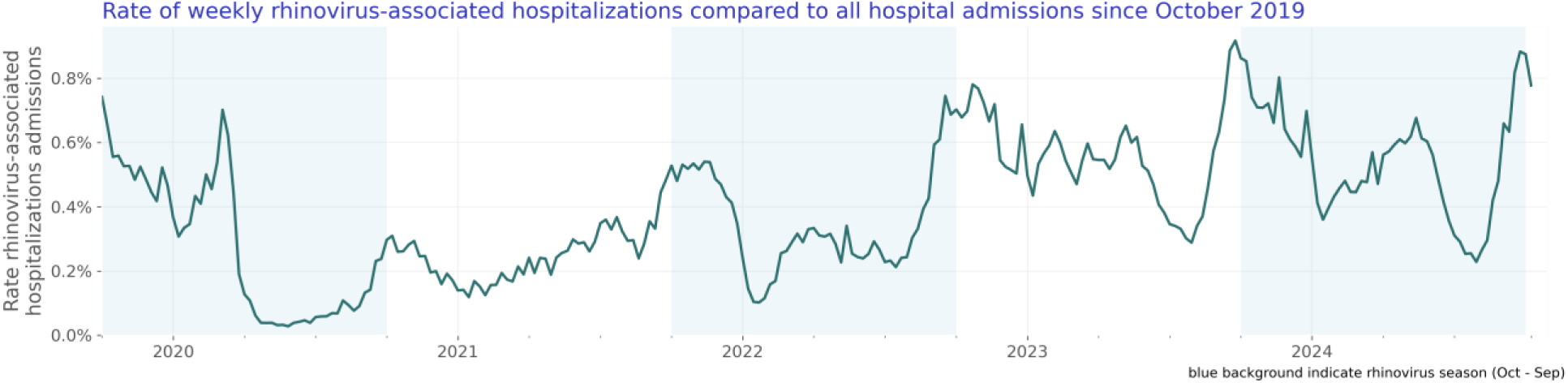
Rate of weekly rhinovirus-associated hospitalizations compared to all hospital admissions since October 2019

**Figure 25:**
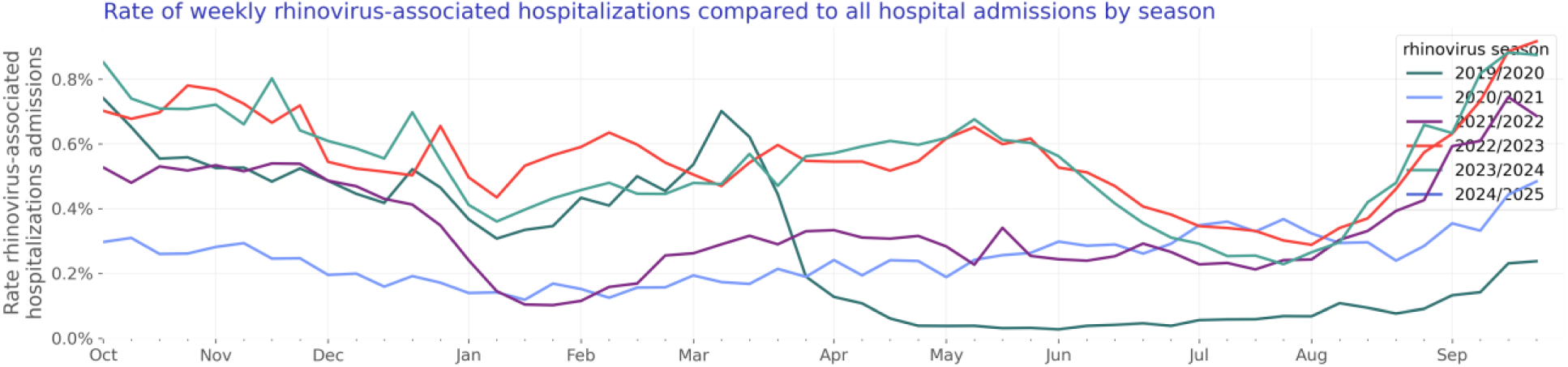
Rate of weekly rhinovirus-associated hospitalizations compared to all hospital admissions by season

#### Test positivity rate over time

We included 1,538,776 rhinovirus lab results with known results of which 236,066 were positive. The rhinovirus test positivity rate is shown in Figure 26. Figure 27 shows yearly trends.

**Figure 26:**
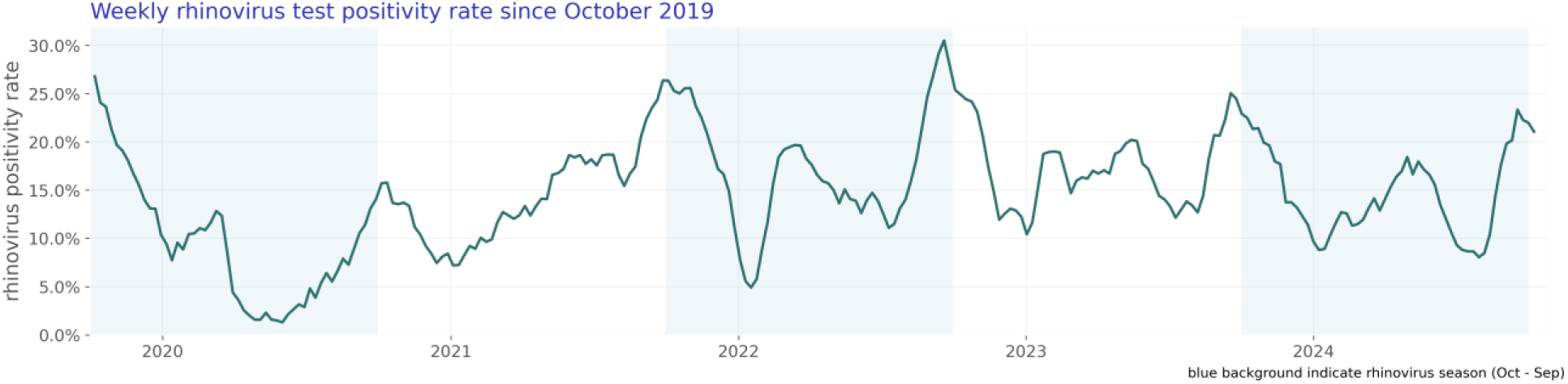
Weekly rhinovirus test positivity rate since October 2019

**Figure 27:**
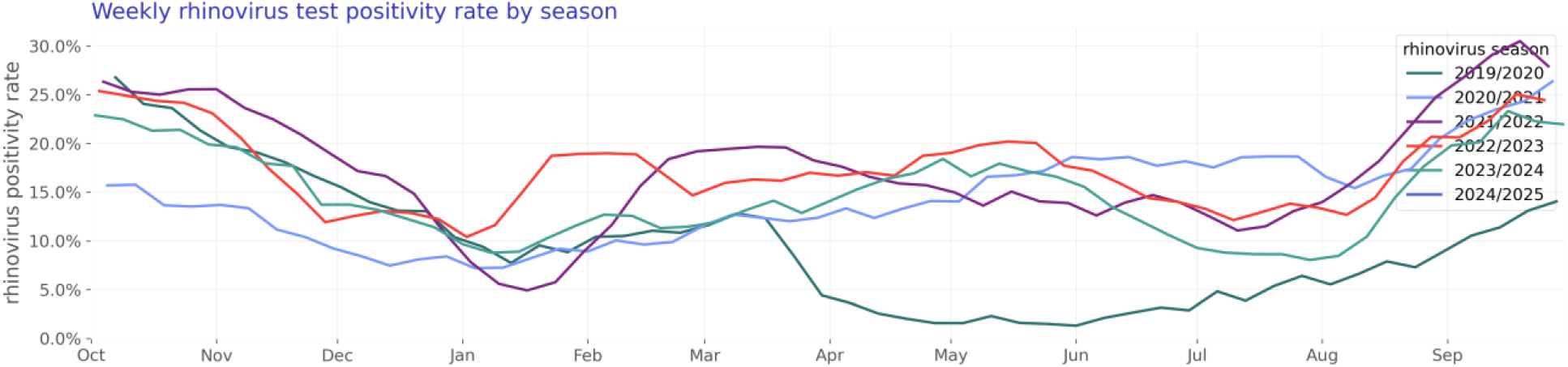
Weekly rhinovirus test positivity rate by season

### Infants and children (age 0-4)

Our infants and children study population consists of 42,903 hospitalizations of 33,336 unique patients (Table 8).

**Table 8:** Infants and children patient characteristics by season.

|  | 2019/2020 | 2020/2021 | 2021/2022 | 2022/2023 | 2023/2024 | Overall |
| --- | --- | --- | --- | --- | --- | --- |
| N | 4,684 (100.00%) | 3,650 (100.00%) | 7,709 (100.00%) | 9,760 (100.00%) | 7,533 (100.00%) | 33,336 (100.00%) |
| Age Group |  |  |  |  |  |  |
| < 6 months | 1,710 (36.51%) | 1,303 (35.70%) | 2,493 (32.34%) | 3,120 (31.97%) | 2,404 (31.91%) | 11,030 (33.09%) |
| 6 - <12 months | 762 (16.27%) | 508 (13.92%) | 1,116 (14.48%) | 1,478 (15.14%) | 1,223 (16.24%) | 5,087 (15.26%) |
| 1 - <2 years | 968 (20.67%) | 867 (23.75%) | 1,693 (21.96%) | 2,042 (20.92%) | 1,703 (22.61%) | 7,273 (21.82%) |
| 2 - 4 years | 1,244 (26.56%) | 972 (26.63%) | 2,407 (31.22%) | 3,120 (31.97%) | 2,203 (29.24%) | 9,946 (29.84%) |
| Sex |  |  |  |  |  |  |
| Female | 2,058 (43.94%) | 1,540 (42.19%) | 3,215 (41.70%) | 4,234 (43.38%) | 3,271 (43.42%) | 14,318 (42.95%) |
| Male | 2,625 (56.04%) | 2,109 (57.78%) | 4,482 (58.14%) | 5,517 (56.53%) | 4,249 (56.41%) | 18,982 (56.94%) |
| Unknown | 1 (0.02%) | 1 (0.03%) | 12 (0.16%) | 9 (0.09%) | 13 (0.17%) | 36 (0.11%) |
| Race |  |  |  |  |  |  |
| White | 2,216 (47.31%) | 1,713 (46.93%) | 3,740 (48.51%) | 4,596 (47.09%) | 3,467 (46.02%) | 15,732 (47.19%) |
| Black or African American | 809 (17.27%) | 714 (19.56%) | 1,289 (16.72%) | 1,498 (15.35%) | 1,182 (15.69%) | 5,492 (16.47%) |
| Asian | 294 (6.28%) | 177 (4.85%) | 505 (6.55%) | 797 (8.17%) | 463 (6.15%) | 2,236 (6.71%) |
| Native Hawaiian or Other Pacific Islander | 85 (1.81%) | 67 (1.84%) | 164 (2.13%) | 205 (2.10%) | 167 (2.22%) | 688 (2.06%) |
| American Indian or Alaska Native | 53 (1.13%) | 33 (0.90%) | 72 (0.93%) | 116 (1.19%) | 59 (0.78%) | 333 (1.00%) |
| Other Race | 784 (16.74%) | 665 (18.22%) | 1,229 (15.94%) | 1,587 (16.26%) | 1,207 (16.02%) | 5,472 (16.41%) |
| Unknown | 443 (9.46%) | 281 (7.70%) | 710 (9.21%) | 961 (9.85%) | 988 (13.12%) | 3,383 (10.15%) |
| Ethnicity |  |  |  |  |  |  |
| Hispanic or Latino | 1,034 (22.08%) | 730 (20.00%) | 1,604 (20.81%) | 2,398 (24.57%) | 1,883 (25.00%) | 7,649 (22.95%) |
| Not Hispanic or Latino | 3,128 (66.78%) | 2,641 (72.36%) | 5,553 (72.03%) | 6,603 (67.65%) | 5,048 (67.01%) | 22,973 (68.91%) |
| Unknown | 522 (11.14%) | 279 (7.64%) | 552 (7.16%) | 759 (7.78%) | 602 (7.99%) | 2,714 (8.14%) |
| Virus |  |  |  |  |  |  |
| COVID | 320 (6.83%) | 601 (16.47%) | 1,743 (22.61%) | 1,114 (11.41%) | 1,154 (15.32%) | 4,932 (14.79%) |
| HMPV | 234 (5.00%) | 23 (0.63%) | 419 (5.44%) | 603 (6.18%) | 353 (4.69%) | 1,632 (4.90%) |
| RSV | 2,002 (42.74%) | 978 (26.79%) | 1,811 (23.49%) | 3,200 (32.79%) | 2,402 (31.89%) | 10,393 (31.18%) |
| influenza | 406 (8.67%) | 4 (0.11%) | 129 (1.67%) | 517 (5.30%) | 414 (5.50%) | 1,470 (4.41%) |
| parainfluenza virus | 240 (5.12%) | 348 (9.53%) | 525 (6.81%) | 781 (8.00%) | 480 (6.37%) | 2,374 (7.12%) |
| rhinovirus | 1,482 (31.64%) | 1,696 (46.47%) | 3,082 (39.98%) | 3,545 (36.32%) | 2,730 (36.24%) | 12,535 (37.60%) |
| Comorbidity |  |  |  |  |  |  |
| Asthma | 357 (7.62%) | 248 (6.79%) | 456 (5.92%) | 636 (6.52%) | 511 (6.78%) | 2,208 (6.62%) |
| Chronic Lung Disease | 50 (1.07%) | 21 (0.58%) | 35 (0.45%) | 62 (0.64%) | 59 (0.78%) | 227 (0.68%) |
| Chronic Obstructive Pulmonary Disease | 10 (0.21%) | 3 (0.08%) | 11 (0.14%) | 13 (0.13%) | 16 (0.21%) | 53 (0.16%) |

#### Hospitalization rate over time

The rate of respiratory virus-associated hospitalizations compared to all hospitalizations for infants and children under five is shown in Figure 28. Figure 29 shows the same data stacked to represent the combined impact of the viruses.

**Figure 28:**
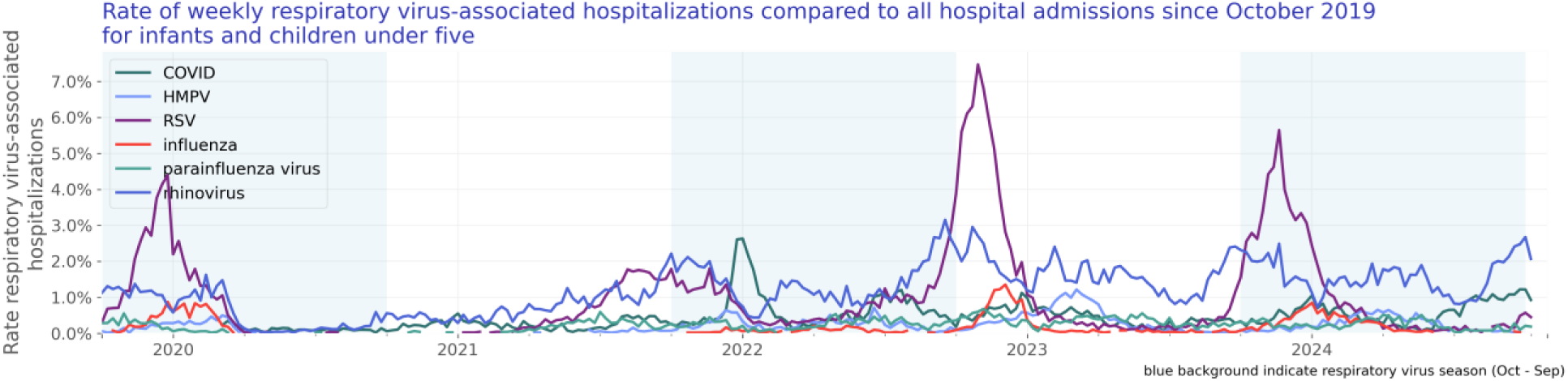
Rate of weekly respiratory virus-associated hospitalizations compared to all hospital admissions since October 2019 for infants and children under five

**Figure 29:**
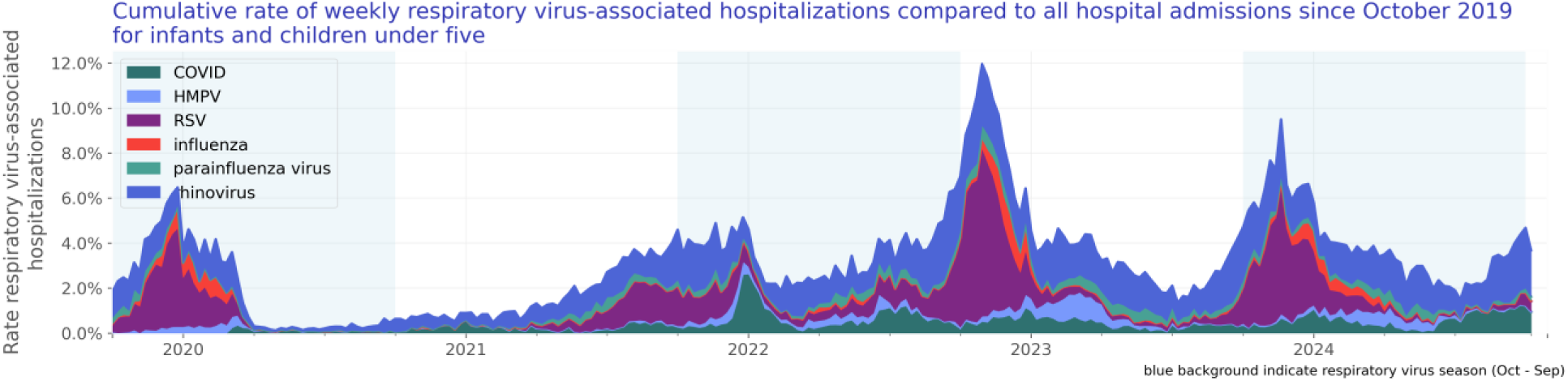
Cumulative rate of weekly respiratory virus-associated hospitalizations compared to all hospital admissions since October 2019 for infants and children under five

#### Test positivity rate over time

We included 5,029,546 lab results with known results of infants and children under age five. Of those tests, 569,165 lab results were positive. The test positivity rate for infants and children under age five is shown in Figure 30.

**Figure 30:**
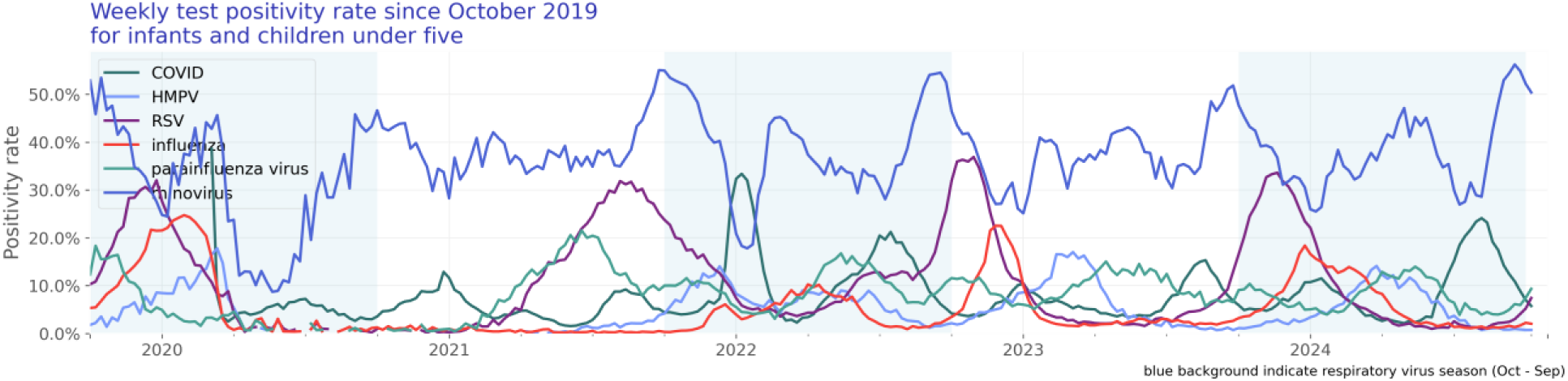
Weekly test positivity rate since October 2019 for infants and children under five

### Older adults (age 65 and over)

Our older adults study population consists of 398,964 hospitalizations of 371,667 unique patients (Table 9).

**Table 9:** Older adults patient characteristics by season.

|  | 2019/2020 | 2020/2021 | 2021/2022 | 2022/2023 | 2023/2024 | Overall |
| --- | --- | --- | --- | --- | --- | --- |
| N | 45,991<br>(100.00%) | 86,736<br>(100.00%) | 93,987<br>(100.00%) | 75,086<br>(100.00%) | 69,867<br>(100.00%) | 371,667<br>(100.00%) |
| Age Group |  |  |  |  |  |  |
| 65 - 74 years | 18,738 (40.74%) | 37,048 (42.71%) | 36,141 (38.45%) | 24,772 (32.99%) | 23,095 (33.06%) | 139,794<br>(37.61%) |
| 75 - 84 years | 16,684 (36.28%) | 31,558 (36.38%) | 35,108 (37.35%) | 29,195 (38.88%) | 27,570 (39.46%) | 140,115<br>(37.70%) |
| 85+ years | 10,569 (22.98%) | 18,130 (20.90%) | 22,738 (24.19%) | 21,119 (28.13%) | 19,202 (27.48%) | 91,758 (24.69%) |
| Sex |  |  |  |  |  |  |
| Female | 23,395 (50.87%) | 42,300 (48.77%) | 47,188 (50.21%) | 39,576 (52.71%) | 36,662 (52.47%) | 189,121<br>(50.88%) |
| Male | 22,397 (48.70%) | 44,145 (50.90%) | 46,470 (49.44%) | 35,193 (46.87%) | 32,731 (46.85%) | 180,936<br>(48.68%) |
| Unknown | 199 (0.43%) | 291 (0.34%) | 329 (0.35%) | 317 (0.42%) | 474 (0.68%) | 1,610 (0.43%) |
| Race |  |  |  |  |  |  |
| White | 29,129 (63.34%) | 62,649 (72.23%) | 70,861 (75.39%) | 56,631 (75.42%) | 51,498 (73.71%) | 270,768<br>(72.85%) |
| Black or African American | 5,403 (11.75%) | 7,391 (8.52%) | 7,542 (8.02%) | 5,570 (7.42%) | 6,171 (8.83%) | 32,077 (8.63%) |
| Asian | 1,065 (2.32%) | 1,582 (1.82%) | 1,791 (1.91%) | 1,978 (2.63%) | 1,697 (2.43%) | 8,113 (2.18%) |
| Native Hawaiian or Other Pacific Islander | 80 (0.17%) | 147 (0.17%) | 191 (0.20%) | 178 (0.24%) | 144 (0.21%) | 740 (0.20%) |
| American Indian or Alaska Native | 133 (0.29%) | 253 (0.29%) | 327 (0.35%) | 327 (0.44%) | 282 (0.40%) | 1,322 (0.36%) |
| Other Race | 2,044 (4.44%) | 3,151 (3.63%) | 2,846 (3.03%) | 2,937 (3.91%) | 2,883 (4.13%) | 13,861 (3.73%) |
| Unknown | 8,137 (17.69%) | 11,563 (13.33%) | 10,429 (11.10%) | 7,465 (9.94%) | 7,192 (10.29%) | 44,786 (12.05%) |
| Ethnicity |  |  |  |  |  |  |
| Hispanic or Latino | 2,723 (5.92%) | 4,567 (5.27%) | 4,067 (4.33%) | 4,185 (5.57%) | 4,668 (6.68%) | 20,210 (5.44%) |
| Not Hispanic or Latino | 33,795 (73.48%) | 67,689 (78.04%) | 77,426 (82.38%) | 62,157 (82.78%) | 58,598 (83.87%) | 299,665<br>(80.63%) |
| Unknown | 9,473 (20.60%) | 14,480 (16.69%) | 12,494 (13.29%) | 8,744 (11.65%) | 6,601 (9.45%) | 51,792 (13.94%) |
| Virus |  |  |  |  |  |  |
| COVID | 30,444 (66.20%) | 83,599 (96.38%) | 85,148 (90.60%) | 52,856 (70.39%) | 44,169 (63.22%) | 296,216<br>(79.70%) |
| HMPV | 1,538 (3.34%) | 28 (0.03%) | 1,067 (1.14%) | 2,158 (2.87%) | 1,901 (2.72%) | 6,692 (1.80%) |
| RSV | 2,777 (6.04%) | 545 (0.63%) | 1,557 (1.66%) | 3,078 (4.10%) | 5,037 (7.21%) | 12,994 (3.50%) |
| influenza | 7,238 (15.74%) | 172 (0.20%) | 2,561 (2.72%) | 9,441 (12.57%) | 11,152 (15.96%) | 30,564 (8.22%) |
| parainfluenza virus | 692 (1.50%) | 563 (0.65%) | 1,035 (1.10%) | 2,148 (2.86%) | 2,048 (2.93%) | 6,486 (1.75%) |
| rhinovirus | 3,302 (7.18%) | 1,829 (2.11%) | 2,619 (2.79%) | 5,405 (7.20%) | 5,560 (7.96%) | 18,715 (5.04%) |
| Comorbidity |  |  |  |  |  |  |
| Asthma | 3,879 (8.43%) | 6,367 (7.34%) | 8,082 (8.60%) | 7,693 (10.25%) | 7,788 (11.15%) | 33,809 (9.10%) |
| Chronic Lung Disease | 3,519 (7.65%) | 6,233 (7.19%) | 7,315 (7.78%) | 6,573 (8.75%) | 6,468 (9.26%) | 30,108 (8.10%) |
| Chronic Obstructive Pulmonary Disease | 8,560 (18.61%) | 14,266 (16.45%) | 19,251 (20.48%) | 16,919 (22.53%) | 16,888 (24.17%) | 75,884 (20.42%) |

#### Hospitalization rate over time

The rate of respiratory virus-associated hospitalizations compared to all hospitalizations for adults 65 and over is shown in ’Figure 31. Figure 32 shows the same data stacked to represent the combined impact of the viruses.

**Figure 31:**
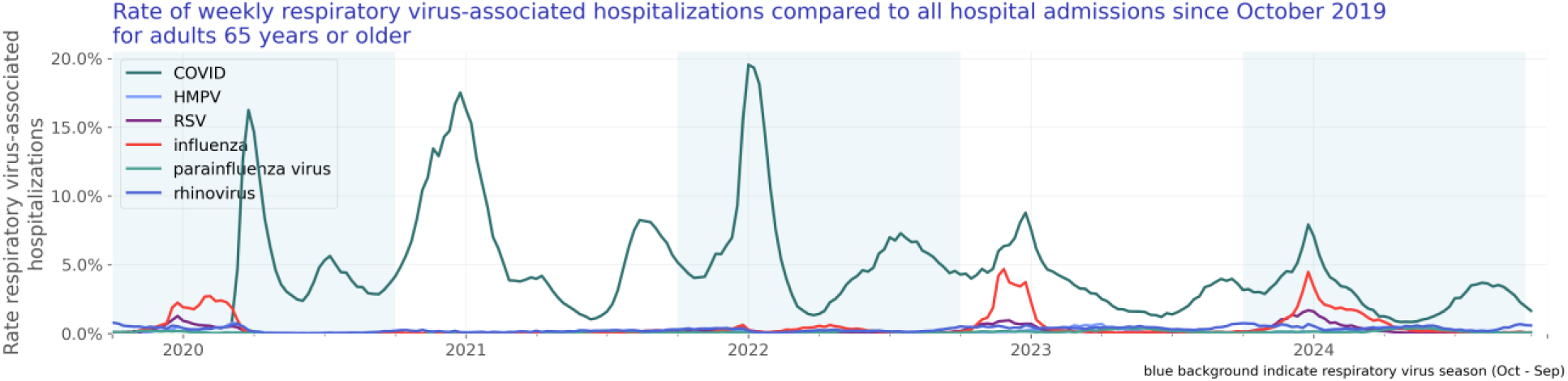
Rate of weekly respiratory virus-associated hospitalizations compared to all hospital admissions since October 2019 for adults 65 years or older

**Figure 32:**
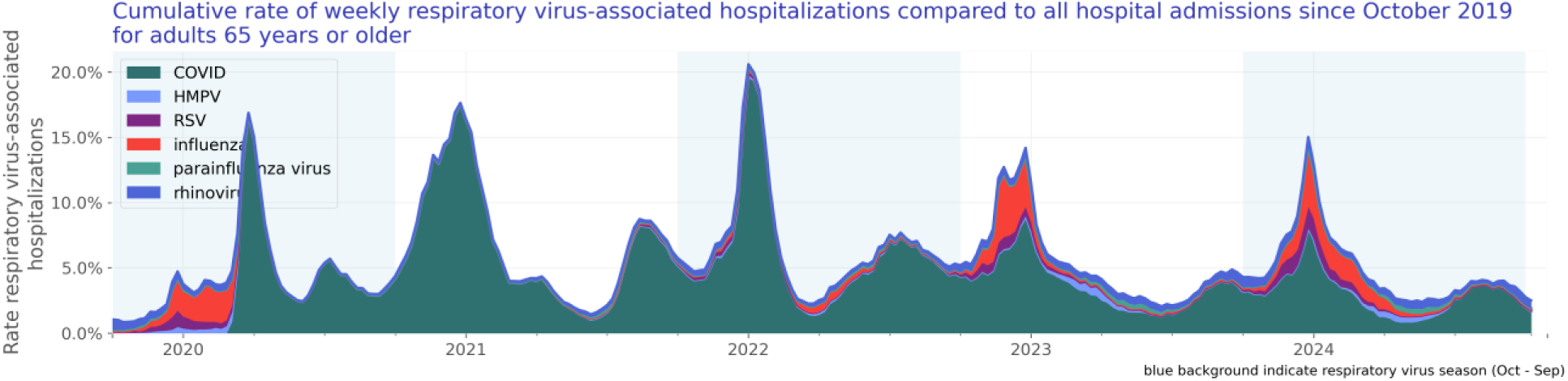
Cumulative rate of weekly respiratory virus-associated hospitalizations compared to all hospital admissions since October 2019 for adults 65 years or older

#### Test positivity rate over time

We included 15,795,812 lab results with known results of adults aged 65 and over. Of those tests, 1,133,937 lab results were positive. The test positivity rate for adults aged 65 and over is shown in Figure 33.

**Figure 33:**
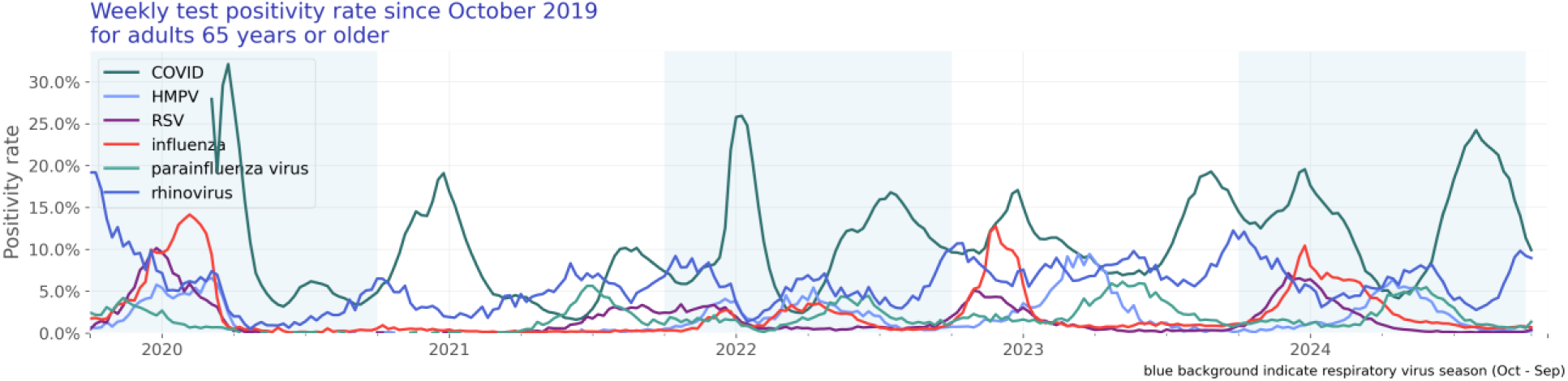
Rate of weekly respiratory virus-associated hospitalizations compared to all hospital admissions since October 2019 for adults 65 years or older

## Discussion and conclusion

Overall, the rate of hospitalizations associated with respiratory viruses decreased by 27.9% in the first week of October 2024 compared to September 2024. The decline in COVID-associated hospitalizations (-46.1%) was substantial, while influenza-associated hospitalizations remained unchanged. Notably, parainfluenza and RSV-associated hospitalizations saw substantial increases of 132.6% and 149.9%, respectively, alongside a smaller increase in rhinovirus-associated hospitalizations (+22.8%). In the first week of October, respiratory virus-associated hospitalizations accounted for 2.0% of all hospitalizations.

In the population aged 0-4 years, respiratory virus-associated hospitalizations increased slightly by 2.9% throughout September 2024. COVID-associated hospitalizations decreased by 14.3%, while parainfluenza-associated hospitalizations rose drastically (+242.9%) and RSV-associated hospitalizations increased by 177.6%. In the first week of October, 2.8% of all hospitalizations in this age group were associated with a respiratory virus, with rhinovirus being the largest contributor at 2.1%.

Among older adults (age 65 and over), there was a decrease of 34.0% in hospitalizations associated with respiratory viruses throughout September 2024. COVID-associated hospitalizations decreased by 46.4%, while influenza-associated hospitalizations remained stable. In the first week of October, respiratory virus-associated hospitalizations accounted for 2.4% of all hospitalizations in this age group, with rhinovirus being a notable contributor at 0.6%.

Several limitations of our analysis should be noted. All data are preliminary and may change as additional data are obtained. These findings are consistent with data accessed October 24, 2024. These are raw counts and post-stratification methods have not been conducted. This analysis does not include patients hospitalized with a respiratory virus who were not tested or were tested later in their medical care (when laboratory tests results would have returned a negative result). In addition, cohorts with small counts may be suppressed during the de-identification process leading to the appearance of zero patients for a given time period. Moreover, the unknowns in this report either indicate the value was not included in the individual’s electronic health record or that it was excluded from the data to protect an individual’s identity as a part of Truveta’s commitment to privacy (Truveta, 2024).

We will continue to monitor respiratory virus-associated hospitalization overall and for the at-risk populations.

## Data Availability

The data used in this study are available to all Truveta subscribers and may be accessed at studio.truveta.com.

